# Effects of Human Lactoferrin (effera^®^) at Two Doses versus Bovine Lactoferrin on the Adult Gut Microbiome and Fecal Short-Chain Fatty Acids: A Randomized, Double-Blind Trial

**DOI:** 10.64898/2025.12.31.25343278

**Authors:** Ross D. Peterson, Julian van der Made, Nicole Kaplan, Sharon M. Donovan, Mei Wang, Ryan N. Dilger, Anthony J. Clark

## Abstract

**Background/Objectives:** Human lactoferrin (hLF) is glycoprotein of commercial interest as a food ingredient for gut health. Here, we report an exploratory analysis evaluating the effects of Helaina hLF (effera^®^), produced by *Komagataella phaffii*, on the adult gut microbiome and fecal metabolites in comparison to bovine LF (bLF).

**Methods:** In a randomized, double-blind, parallel-arm, controlled trial, 66 healthy adults received either high-dose (HD) effera^®^ (3.4 g/day), low-dose (LD) effera^®^ (0.34 g/day), or bLF (3.4 g/day) supplementation for 28 days. Fecal samples were collected at baseline (Day 0), Day 28, Day 56, and Day 84 and analyzed for microbial diversity, taxonomic shifts, and volatile fatty acids (VFA).

**Results:** Alpha-diversity remained stable across all groups. Beta-diversity showed no main effect of treatment; however, bLF was associated with significant visit-related shifts, as assessed by weighted UniFrac. At the phylum level, significant changes associated with effera^®^ were observed, including decreases in Bacillota (LD) and Verrucomicrobiota (HD), and notable genera increases in *Lachnospira*, *Paraprevotella*, and *Faecalibacterium* (HD), while bLF was associated with an increase in *Roseburia*. Both effera^®^ and bLF were associated with decreases in *Blautia* and *Dorea*. VFA analysis revealed that bLF increased absolute total short-chain fatty acids (SCFAs) and branched-chain fatty acids (BCFAs) concentrations, while both effera^®^ groups produced proportional changes in SCFAs, individual BCFAs, and acetate.

**Conclusions:** In healthy adults, effera^®^ supplementation promoted a proportional increase in acetate and supported potentially beneficial taxa while maintaining microbial diversity, without disrupting community structure. (clinicaltrials.gov: NCT06012669).

## 1. Introduction

Human lactoferrin (hLF) is a single polypeptide chain of approximately 80 kDa, consisting of around 690–691 amino acid residues with three glycosylation sites.^1^ The tertiary structure of hLF is characterized by two homologous globular domains, the N- and C-terminal lobes, which consist of α-helices, β-pleated sheets, and turns.^2^ Each lobe contains a deep cleft capable of binding one ferric iron ion.^3^ hLF has a reversible iron-binding property that gives rise to three forms: iron-depleted (apoLF), a monoferric form, and iron-saturated (holoLF).^3^ LF has been shown to play a role in various physiological processes, including support of gut barrier function,^4,5^ immune development,^6^ iron homeostasis,^7^ and the gut microbiome.^8,9^

LF exerts multifaceted effects to influence the gut microbiome.^10^ The bacteriostatic effects of LF can affect the growth of pathogenic bacteria, including *Escherichia coli*, *Salmonella spp.*, and *Staphylococcus aureus*. In addition, bovine lactoferrin (bLF) has been shown to support beneficial taxa, such as *Bifidobacterium* and *Lactobacillus* spp.^11,12^ The growth-promoting activity of LF can be strain-dependent and influenced by the degree of iron saturation. For example, holo-bLF enhances the growth of *Lactobacillus acidophilus*, whereas the bifidogenic activity appears less dependent on saturation state.^11^ However, studies show that bLF and hLF each support distinct *Bifidobacterium* strains.^12^ Moreover, proteolytic fragments of LF, such as lactoferricin, not only display direct antimicrobial effects but also can contribute to the enrichment of beneficial taxa.^13^ In one study, holo-bLF supported the growth of commensals, such as *Alistipes putredinis*.^8^ In vivo animal models have further confirmed LF’s ability to shape microbial ecology, with supplementation increasing bacterial richness and the abundance of *Bifidobacterium* and *Lactobacillus spp*., while reducing opportunistic pathogens, including *Escherichia-Shigella*, *Veillonella*, and *Leptotrichia*.^14,15^

In neonates, higher LF levels in breast milk have correlated positively with *Bifidobacterium* and *Lactobacillus* abundance.^16^ These genera have been known to be adapted to low-iron environments, and due to the high abundance of these genera, acetate has been the primary short-chain fatty acid (SCFA) reported in infant stool.^17^ Recombinant hLF (talactoferrin) was shown to modulate *Enterobacteriaceae* in very-low-birth-weight infants.^10^ However, the overall clinical impact on infant gut microbiota has been modest, with some studies reporting only small reductions in opportunistic pathogens, such as *Staphylococcus*, and no significant shifts in diversity.^18,19^ Despite compelling preclinical evidence and modest effects observed in infants, clinical studies directly evaluating LF’s impact on the adult gut microbiome remain scarce, representing a key gap in understanding its translational relevance to adult populations. Notably, early-life LF biology also offers a mechanistic frame for interpreting LF-associated microbiome signals detected in adults.^20^ LF exposure in early life is both quantitatively substantial and developmentally patterned, with concentrations highest in colostrum and declining across lactation, consistent with a role in shaping microbial succession during a period of heightened ecological plasticity.^20^

LF can also modulate epithelial and immune programs, thereby altering the selective pressures that govern colonization at the mucosal interface.^21^ In parallel, infant-adapted *Bifidobacteria* encode enzymes that release and metabolize complex N-glycans from human milk glycoproteins, supporting a selection in the milk-fed intestine.^22^ Consistent with this framework, in a cohort of breastfed mother–infant pairs that included preterm infants, higher early fecal LF concentrations were positively associated with fecal *Bifidobacterium* and *Lactobacillus*, consistent with a potential role for luminal LF in supporting the establishment of beneficial early-life taxa.^23^ Together, these considerations support evaluating whether recombinant hLF preparations can elicit functionally coherent yet non-disruptive microbiome responses in adults, which may be informative for future work across life stages.

Due to the significant interest in LF supplementation, alternative sources beyond animal milk have been developed, including human-equivalent LF alpha. Helaina human lactoferrin (effera^®^) is expressed in the yeast, *Komagataella phaffii* (*K. phaffii*), and is produced at a commercial scale. Helaina effera^®^ possesses an amino acid sequence identical to native human milk lactoferrin (hmLF),^1^ and its secondary structure, as revealed by microfluidic modulation spectroscopy, mirrors that of hmLF with globular structures closely akin to hmLF.^1^ Glycosylation of effera^®^ is consistent with its production in *K. phaffii* and contains N-linked glycans that are predominantly oligomannose structures (M5–M9), which have also been detected in the human milk glycobiome^24^ and in bLF.^25^ Low levels of oligomannose glycans have been detected at the same known glycosylation sites as those of hmLF.^1^ Helaina effera^®^ has been developed as a food ingredient and dietary supplement and has met the Generally Recognized As Safe (GRAS) standard of general acceptance and general availability supported by safety studies in adults.^1,26–31^

bLF has been manufactured on an industrial scale and is widely used in consumer products, such as infant formula, fortified yogurts, and nutritional supplements. In the United States, bLF was granted numerous letters of no objection from the FDA regarding its GRAS status for various intended uses, including infant formula, functional foods, ice cream, and powdered milk, among others.^32–36^ The existing literature highlights the beneficial effects of LF on the gut, but comprehensive evidence on its impact on the healthy adult gut remains limited, with most research focusing on preclinical models or vulnerable populations (e.g., preterm infants).^10,18,27^

To our knowledge, no study has evaluated the impact of orally consumed fermentation-derived hLF on the healthy adult gut microbiome. The current study presents exploratory analyses of the fecal microbiota composition and volatile fatty acid (VFA) nested within a randomized, double-blind, controlled, parallel-arm trial. The hypothesis-generating analysis reported herein investigated shifts in alpha- and beta-diversity of the microbiome, microbial composition, and concentrations of VFA in fecal samples collected at multiple time points.

## 2. Materials and Methods

### 2.1. Sample collection

The original study, which enrolled healthy adults (18–45 years), was designed to assess the safety and immunogenicity/alloimmunization potential of Helaina effera^®^.^27^ Participants in the original study received one of three treatments: high-dose (HD) effera^®^ (3.4 g/d, 1.7 g twice daily), low-dose (LD) effera^®^ (0.34 g/d, 0.17 g twice daily), or bLF as the active control (3.4 g/d, 1.7 g twice daily) over a 28-day dietary intervention period.^27^ Helaina effera^®^ had an iron saturation of ∼50%, whereas bLF exhibited <10% saturation. Fecal samples (n = 195) were collected at four time points during the clinical study (Figure 1, Table 1). Participants were provided with a sample collection kit and instructed to collect a freshly voided fecal sample during the 72 hours prior to the baseline visit (Day 0) and 24 hours prior to their subsequent visits on Day 28, Day 56, and Day 84. Results of additional analyses of microbiome data from follow-up visits (i.e., Day 56 and Day 84) are available as supplementary figures and tables. All samples were placed in separate sterile tubes, and frozen at -20 °C until delivered to the clinical site during a clinical visit. The samples were immediately placed in a -80 °C freezer for storage until shipped on dry ice to the University of Illinois for analysis. The primary analysis presented in this paper compares baseline (Day 0) to Day 28 across the three treatments.

**Figure 1.**
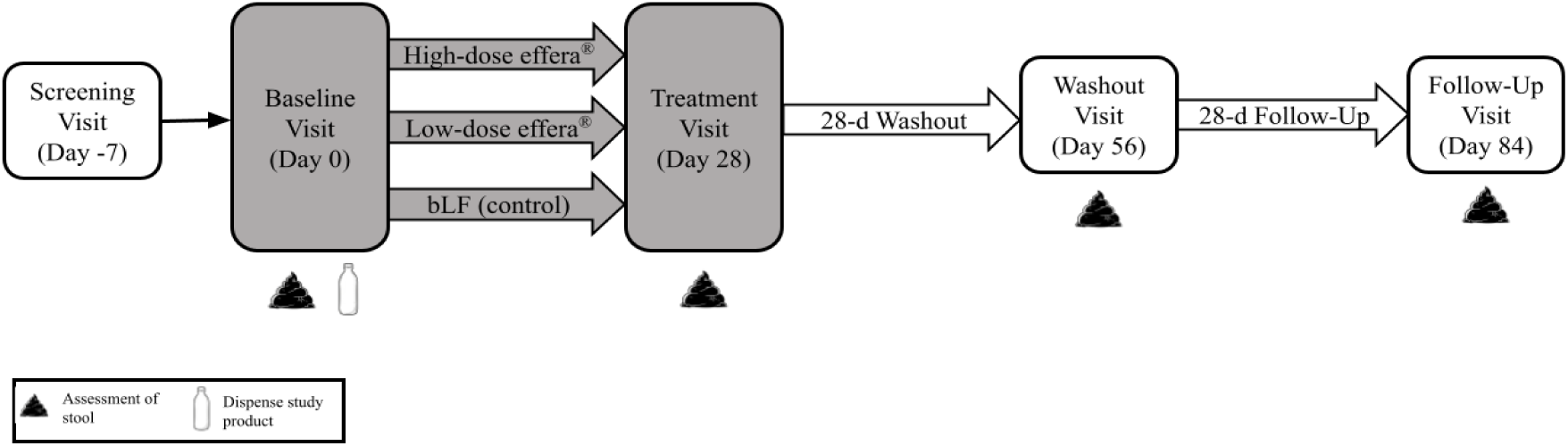
Study design schematic for microbiome sample fecal collection. Assessments on fecal microbiome included alpha- and beta-diversity, taxonomy, and VFA concentrations, with comparisons between baseline (Day 0) and treatment visit (Day 28). Abbreviation: bLF, bovine lactoferrin. Grayed boxes indicate the treatment phase of the study.

**Table 1.** Number of fecal samples collected from healthy adults at baseline, treatment visit, washout visit, and follow-up visit.

| Time point | Number of samples |
| --- | --- |
| Baseline visit, Day 0 | 56 |
| Treatment visit, Day 28 | 49 |
| Washout visit, Day 56 | 45 |
| Follow-up visit, Day 84 | 45 |

### 2.2. DNA isolation

DNA was extracted from ∼200 mg of feces using the QIAamp Fast DNA Stool Mini Kit (Qiagen, Valencia, CA) in combination with bead beating on the FastPrep-24 System (MP Biomedicals, Carlsbad, CA) as previously described.^37^ DNA concentration was measured on a Qubit 3.0 Fluorometer using the Qubit 1X dsDNA HS Assay Kit (Thermo Fisher Scientific, Waltham, MA), and its quality was checked on a 1% agarose gel.

### 2.3. Library preparation and sequencing of full-length 16S rRNA genes

Construction of full-length 16S rRNA gene libraries and sequencing on the PacBio REVIO were performed at the Roy J. Carver Biotechnology Center, University of Illinois at Urbana-Champaign. The full-length 16S rRNA amplicons were generated from 2.5ng of DNA with the barcoded Kinnex primers (forward primer: 5’-AGRGTTYGATYMTGGCTCAG-3’ and the reverse primer: 5’-RGYTACCTTGTTACGACTT-3’) following the protocol from PacBio (PacBio, Menlo Park, CA). The individually barcoded amplicons were pooled and converted to a library with the SMRTBell Template Prep kit 3.0 (PacBio). The library was quantified using a Qubit 3.0 (Thermo Fisher Scientific) and run on a Fragment Analyzer (Agilent Technologies, Santa Clara, CA) to confirm the presence of DNA fragments of the expected size. The library was sequenced with SPRQ chemistry on a SMRT cell in the PacBio REVIO instrument with a 30-hour movie time. Circular consensus sequence (CCS) analysis and demultiplexing were performed using SMRTlink 25.1, applying the following parameters: ccs --min-passes 3 --min-rq 0.999.

### 2.4. Sequence processing

Sequences were processed using the DADA2 R package and the QIIME 2 pipeline.^38,39^ Demultiplexed reads obtained from the sequencing facility were primer removed, dereplicated, denoised, and chimera checked using DADA2 (version 1.36.0).^38^ The amplicon sequence variant (ASV) table and representative sequences generated from DADA2 were imported and further analyzed in QIIME 2 (version 2025.4). The representative sequences were aligned, and a phylogenetic tree was constructed from the filtered alignment as previously described.^40^ Alpha-diversity (observed features, Shannon entropy, Pielou’s evenness, and Faith’s phylogenetic diversity) and UniFrac distance metrics were computed using the plugin diversity with the core metrics-phylogenetic method on the ASV table and phylogenetic tree. Samples were rarefied to an equal number of reads (13,900) for calculating alpha-diversity and UniFrac distance metrics. Taxonomic assignments were performed using the plugin feature-classifier with the classify sklearn method,^39^ in which a pre-trained Naïve-Bayes classifier with Greengenes2 2024.09 full-length sequences was used (https://library.qiime2.org/data-resources).

Taxonomic bar plots were generated using the plugin taxa with the taxa barplot method. The bar plots were visualized using QIMME 2 View,^41^ and the count tables at the phylum and genus levels were downloaded for differential abundance analysis.

### 2.5. Volatile fatty acid concentrations

The concentrations of SCFA (acetate, butyrate, and propionate) and branch-chain fatty acids (BCFA; isobutyrate, isovalerate, and valerate) were analyzed by gas chromatography (model 5890A series 99; Hewlett-Packard, Palo Alto, CA) using a glass capillary column (100 m × 0.25 mm i.d. × 0.2 µm film thickness; model SP-2560; Supelco, Bellefonte, PA).

Oven temperature, detector temperature, and injector temperature were 125 °C, 175 °C, and 180°C, respectively. All VFA concentrations (μmol/g) were expressed on a dry matter basis.

### 2.6. Statistical analysis

Alpha-diversity was analyzed using the lmer function of the lme4 R package (version 4.5.1).^42^ The effect of treatment and visit on beta-diversity was evaluated with principal coordinate analysis (PCoA) and permutational multivariate analysis of variance (PERMANOVA) based on UniFrac distances. The statistical model included treatment, visit, and the interaction between treatment and visit as fixed effects, and participant identification as a random effect. Total fiber intake was included in the model as a covariate. PCoA plots were generated using QIIME2. PERMANOVA was performed using the adonis2 function of the R package vegan 2.7-1.QIIME2 plugin q2-longitudinal (pairwise-distances method) was applied to compare the within-subject distance between baseline visit (Day 0) and Day 28 for each treatment.^42^ Statistical significance was set at *P* ≤ 0.05.

Multivariate analysis by linear models (MaAsLin 3.0)^43^ was used to determine associations between visit and the relative abundance of bacterial taxa within each treatment. The relative abundance data were log2-transformed; only the taxa with relative abundances ≥ 0.1% in more than 10% of the samples were included in the analyses. All *P*-values were corrected for multiple testing by the Benjamini-Hochberg false discovery rate (FDR), and *P*-values ≤ 0.05 were considered statistically significant. And 0.05 < *P* ≤ 0.10 were reported as trends toward significance.

The proportions of SCFA and BCFA in total fatty acids were calculated from gas chromatography data for each participant at each study visit. Additionally, the proportion of each fatty acid was first expressed as a fraction of its respective class (SCFA or BCFA). Data from the baseline visit (Day 0), the end of the 28-day dietary intervention period, and follow-up visits at Days 56 and 84 were used in the analysis. Participants with any missing data were removed, and one participant from the HD effera^®^ group was excluded due to an outlier value of butyrate in the 28-day sample. Due to the high degree of dietary record variability, no covariates (e.g., fiber) were used in the analysis.

For each participant, the percentage change in VFA proportion relative to the baseline visit was computed for each metabolite. Group-level summary statistics were then calculated by visit, protocol population, and metabolite. Specifically, the median, 25th and 75th percentiles, and standard deviation of both raw proportions and percent changes were computed.

To assess whether post-treatment changes differed significantly from zero, a Wilcoxon signed-rank test was performed on the percentage change values within each treatment group and metabolite, excluding missing data. To evaluate whether the magnitude of percent change differed significantly between treatment groups, Mann–Whitney U tests were conducted pairwise between protocol populations for each metabolite and visit. Resulting *P*-values were merged into the summary table for each visit and metabolite. All analyses were performed in Python using pandas and scipy.stats libraries.

## 3. Results

### 3.1. Microbiome alpha and beta-diversity measures

#### 3.1.1. Alpha-diversity measures

The alpha-diversity measures of observed features, Shannon entropy, evenness, and Faith’s phylogenetic diversity (Supplementary Table A1) remained stable across treatments and visits, and no interaction effects were identified. These results demonstrate that both HD and LD effera^®^ supplementation maintained the microbiome diversity of healthy adults after the 28-day dietary intervention period.

#### 3.1.2. Beta-diversity measures

The QIIME2 plugin q2-longitudinal (pairwise-distances method) was applied to compare the within-subject distances between baseline (Day 0) and Day 28 for each treatment (Figure 2). When weighted UniFrac distances were used, within-subject distances between the two visits were similar among the treatment groups (*P* > 0.05) (Figure 2A). Based on unweighted Unifrac distances, HD effera^®^ had a smaller within-subject distance than bLF or LD effera^®^ treatments (*P* = 0.009 and *P* = 0.005, respectively) (Figure 2B).

**Figure 2.**
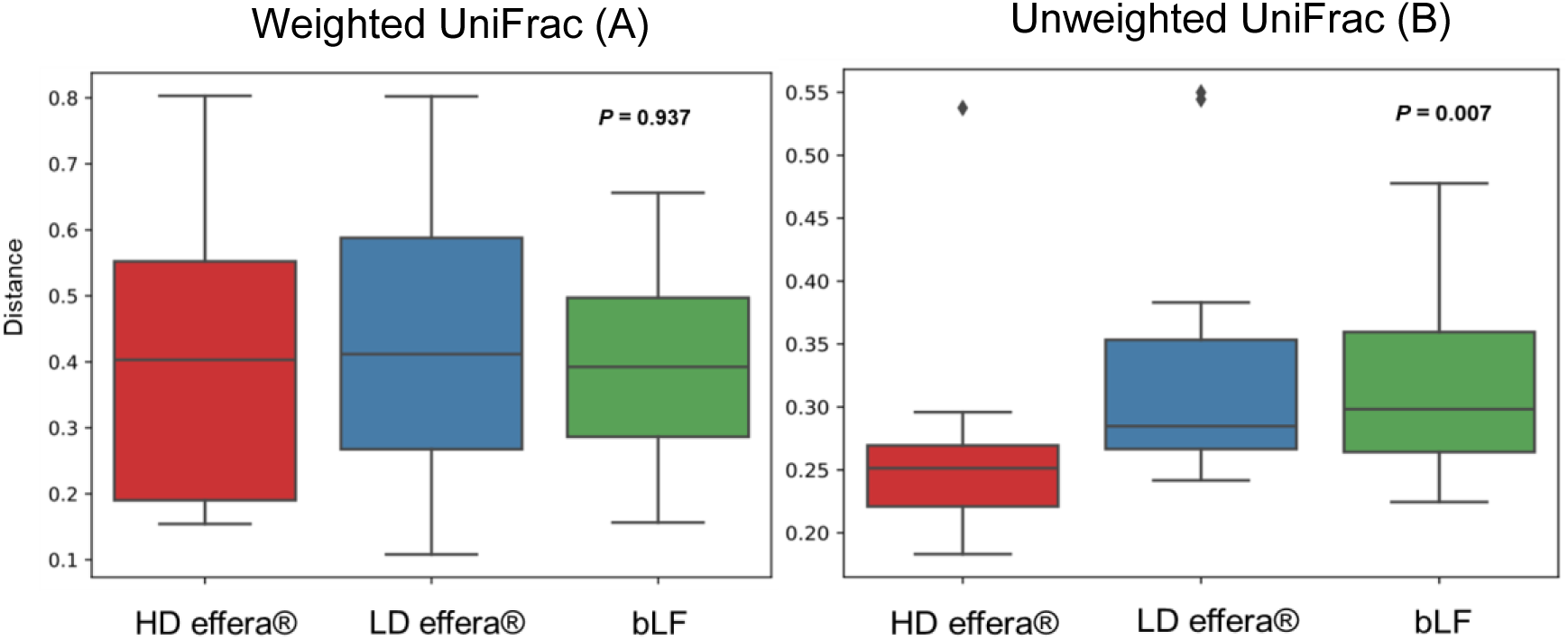
Effects of lactoferrin supplementation on beta-diversity. Within-subject comparisons of weighted (A) and unweighted (B) UniFrac distances between Day 0 to Day 28 across treatment groups were evaluated. Samples were collected from each participant at two time points: baseline (Day 0) and Day 28. The total participants analyzed for HD effera^®^ (3.4 g/d) were n=14, LD effera^®^ (0.34 g/d) were n=15, bLF (3.4 g/d) were n=14. Abbreviations: HD, high-dose effera^®^; LD, low-dose effera^®^; bLF, bovine lactoferrin.

PERMANOVA indicated that neither treatment nor interaction between the treatment and visit impacted beta-diversity (*P* > 0.05 for both weighted and unweighted UniFrac distances). However, there was a global effect of visit based on both weighted and unweighted UniFrac distances (*P* = 0.011 and *P* = 0.020, respectively). To determine which treatment contributed to the visit effect between baseline (Day 0) and the end of the 28-day intervention (Day 28), data were stratified by treatment. bLF supplementation affected visit-related shifts in beta-diversity based on weighted UniFrac distances (*P* = 0.027) but not on unweighted UniFrac distances (*P* > 0.05). There were no visit effects for both the HD and LD effera^®^ treatments (*P* > 0.05).

### 3.2. Microbial compositional shifts in response to effera^®^ and bLF interventions

#### 3.2.1. Phylum Level

Shifts in microbiome taxonomy were observed at the phylum level for bLF and HD and LD effera^®^. Taxonomic stacked bar plots identified various phyla affected by LF supplementation (Figure 3). *Bacteroidota* had a trend increase in median relative abundance in the bLF group (Table 2). In response to LD effera^®^, *Bacillota_I* significantly decreased and Verrucomicrobiota significantly decreased in the HD effera^®^ (Table 2). Taxonomic stacked bar plots for each treatment showing the mean phyla relative abundance from baseline (Day 0) through Day 84 may be found in Supplementary Figure A1.

**Figure 3.**
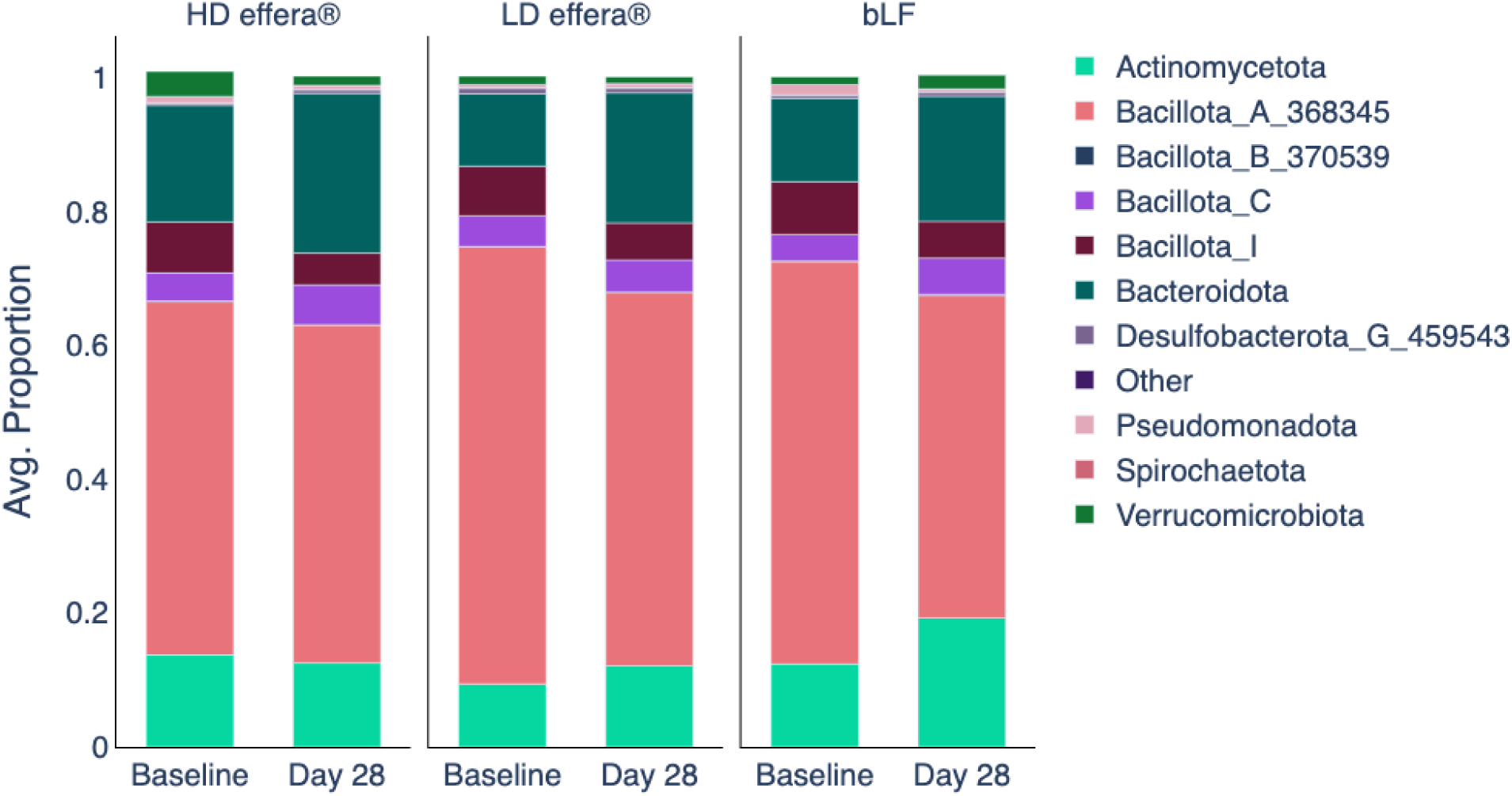
Taxonomic stacked bar plots showing the mean relative abundance of phyla at baseline through supplementation (Day 0 and Day 28) for each treatment group. Counts were normalized to their library size; the 10 most abundant phyla were plotted; “Other” phyla included all remaining phyla not in the top 10. Average proportions were individual relative abundances, averaged across the treatment group. The total participants analyzed for HD effera^®^ (3.4 g/d) were n=14, LD effera^®^ (0.34 g/d) were n=16, bLF (3.4 g/d) were n=15. Abbreviations: HD, high-dose effera®; LD, low-dose effera®; bLF, bovine lactoferrin.

**Table 2.**
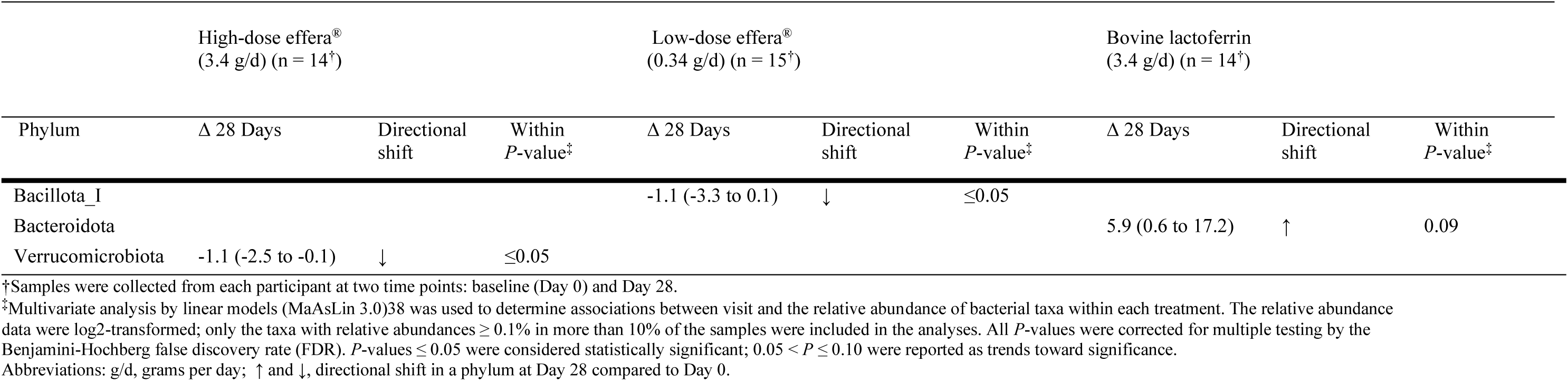
Directional shifts in phyla from baseline (Day 0) to Day 28 in response to high-dose effera^®^ (3.4 g/d), low-dose effera^®^ (0.34 g/d), and bovine lactoferrin (3.4 g/d) treatments in healthy adults. Data are presented as median (25th percentile to 75th percentile).

#### 3.2.2. Genus level

Taxonomic stacked bar plots visually show various genera affected by LF supplementation (Figure 4). In response to the 28-day dietary intervention, several predominant and low-abundance gut bacterial genera exhibited significant, or trend shifts in relative abundance in response to LF supplementation (Table 3). Many shifts were consistent across multiple treatment groups. Exclusive to both effera^®^ groups, significant increases were observed in *Lachnospira, Paraprevotella,* and *Sutterella.* Specific to both the LD effera^®^ and bLF groups significant or trend decreases were measured in *Dorea* (*Dorea_A*), Blautia (*Blautia_A_141781*), and *Fimenecus*; *Dysosmobacter* showed a trend increase in both groups. *Prevotella* was significantly increased in the HD effera^®^ and bLF groups; *Ruminococcus_B* significantly decreased in both groups. Taxonomic stacked bar plots for each treatment showing the mean genera relative abundance from baseline (Day 0) through Day 84 may be found in Supplementary Figure A2.

Certain microbial changes were treatment-group specific. LD effera^®^ produced significant or trend reduction in *Fusicatenibacter, Howardella, Intestinibacter, Limiplasma, Mogibacterium_A,Turicibacter* and increases in *Acidaminococcus, Adlercreutzia_A_404257 Coprococcus (Coprococcus_A_121497), Eubacterium (Eubacterium_F), G11, Lawsonibacter, Parasutterella_564865, Porcincola, UBA6382,* and *Wujia*. The HD effera^®^ was associated with decreases in *Akkermansia, Angelakisella, Clostridium (Clostridium_AP_143938),* and *Ruthenibacterium;* increases were measured in *Agathobacter (Agathobacter_164117), Alistipes (Alistipes_A_871404), Allisonella, Bilophila, Butyricimonas Catenibacterium, Enterocloster,* and *Faecalibacterium*. bLF supplementation led to a significant or trend reduction in relative abundance of *Anaerobutyricum*, *Clostridium* (*Clostridium_AQ and _T*), *Dorea* (*Dorea_D_146760) Eubacterium (Eubacterium_J), Lachnoclostridium (Lachnoclostridium_B) Fusicatenibacter, Limisoma, PeH17, Romboutsia (Romboutsia_B), Schaedlerella* and *Terrisporobacter.* Conversely, bLF significantly increased *Alloprevotella, Bacteroides (Bacteroides_H_857956), Clostridium (Clostridium_A_51961), Cryptobacteroides, Eisenbergiella, Hominicoprocola, Megasphaera_A_38685,* and *Roseburia (Roseburia_C)*.

**Figure 4.**
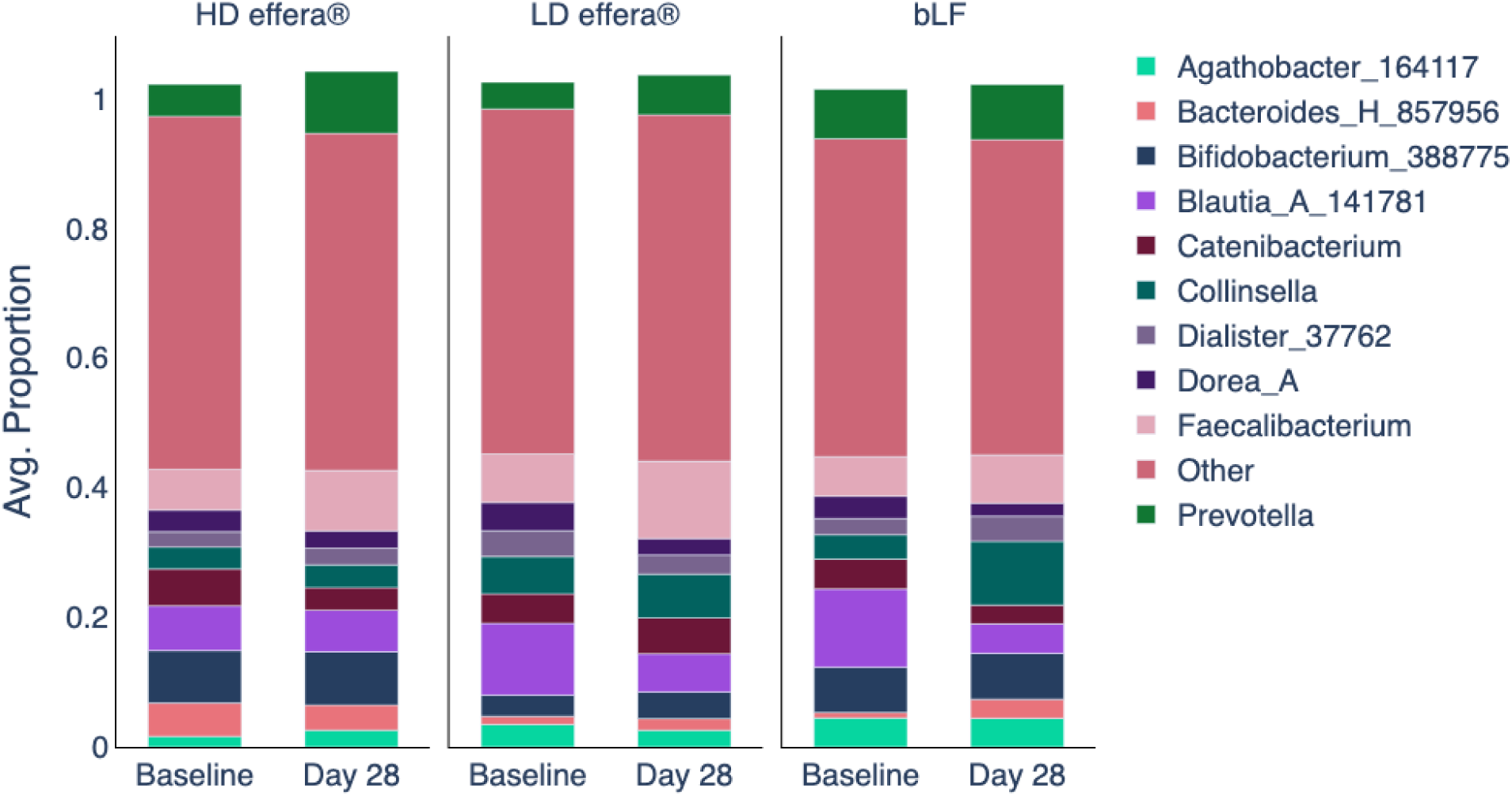
Taxonomic stacked bar plots showing the mean relative abundance of genera at baseline through supplementation (Day 0 and Day 28) for each treatment group. Counts were normalized to their library size; the 10 most abundant genera were plotted; “Other” genera included all remaining genera not in the top 10. Average proportions were individual relative abundances, averaged across the treatment group. The total participants analyzed for HD effera^®^ (3.4 g/d) were n=14, LD effera^®^ (0.34 g/d) were n=16, bLF (3.4 g/d) were n=15. Abbreviations: HD, high-dose effera®; LD, low-dose effera®; bLF, bovine lactoferrin.

**Table 3.** Directional shifts in genera in response to high-dose effera^®^ (3.4 g/d), low-dose effera^®^ (0.34 g/d), and bovine lactoferrin (3.4 g/d) treatments in healthy adults. Data are presented as median (25th percentile to 75th percentile).

| Genus | High-dose effera®<br>(3.4 g/d) (n=14<sup>†</sup>) |  |  | Low-dose effera®<br>(0.34 g/d) (n=15<sup>†</sup>) |  |  | Bovine lactoferrin<br>(3.4 g/d) (n=14<sup>†</sup>) |  |  |
| --- | --- | --- | --- | --- | --- | --- | --- | --- | --- |
|  | Δ 28 Days* | Directional shift | Within P-value<sup>‡</sup> | Δ 28 Days* | Directional shift | Within P-value<sup>‡</sup> | Δ 28 Days* | Directional shift | Within P-value<sup>‡</sup> |
| *Acidaminococcus* |  |  |  | 0.8 (0.2 to 1) | ↑ | 0.06 |  |  |  |
| *Adlercreutzia_A_404257* |  |  |  | 0.1 (0 to 0.6) | ↑ | 0.06 |  |  |  |
| *Agathobacter_164117* | 0 (-0.6 to 3) | ↑ | ≤0.05 |  |  |  |  |  |  |
| *Akkermansia* | -1.4 (-5.8 to -0.8) | ↓ | ≤0.05 |  |  |  |  |  |  |
| *Alistipes_A_871404* | 0.04 (0 to 0.3) | ↑ | ≤0.05 |  |  |  |  |  |  |
| *Allisonella* | 0.02 (0.01 to 0.03) | ↑ | ≤0.05 |  |  |  |  |  |  |
| *Alloprevotella* |  |  |  |  |  |  | 2.1 (0.6 to 3.3) | ↑ | 0.07 |
| *Anaerobutyricum* |  |  |  |  |  |  | -0.3 (-0.9 to 0.2) | ↓ | 0.09 |
| *Anaerostipes* |  |  |  |  |  |  | -0.3 (-0.9 to 0) | ↓ | 0.1 |
| *Angelakisella* | -0.1 (-0.9 to 0) | ↓ | ≤0.05 |  |  |  |  |  |  |
| *Bacteroides_H_857956* |  |  |  |  |  |  | 0.2 (0 to 1.3) | ↑ | 0.1 |
| *Bilophila* | 0.1 (0 to 0.3) | ↑ | 0.1 |  |  |  |  |  |  |
| *Blautia_A_141781* |  |  |  | -8 (-10.4 to 1.7) | ↓ | ≤0.05 | -2.8 (-5.7 to -0.8) | ↓ | ≤0.05 |
| *Butyricimonas* | 0.1 (0 to 0.1) | ↑ | ≤0.05 |  |  |  |  |  |  |
| *Catenibacterium* | -0.3 (-0.6 to 1.7) | ↓ | 0.1 |  |  |  | -0.6 (-2.9 to 1.9) | ↓ |  |
| *Clostridium_A_51961* |  |  |  |  |  |  | 0.1 (0 to 0.2) | ↑ | ≤0.05 |
| *Clostridium_AP_143938* | -0.1 (-0.2 to -0.1) | ↓ | 0.1 |  |  |  |  |  |  |
| *Clostridium_AQ* |  |  |  |  |  |  | -0.1 (-0.4 to 0) | ↓ | 0.08 |
| *Clostridium_T* |  |  |  |  |  |  | -0.2 (-0.9 to 0.5) | ↓ | 0.06 |
| *Coprococcus_A_121497* |  |  |  | 0.1 (0.1 to 0.4) | ↑ | 0.09 |  |  |  |
| *Cryptobacteroides* |  |  |  |  |  |  | 0.6 (0.1 to 1.2) | ↑ | ≤0.05 |
| *Dorea_A* |  |  |  | -2.1 (-3.4 to 0.5) | ↓ | 0.06 | -2 (-3 to -0.4) | ↓ | ≤0.05 |
| *Dorea_D_146760* |  |  |  |  |  |  | -0.2 (-0.4 to 0) | ↓ | 0.06 |
| *Dysosmobacter* |  |  |  | 0 (0 to 0.8) | ↑ | 0.1 | 0.1 (0 to 0.2) | ↑ | 0.1 |
| *Eisenbergiella* |  |  |  |  |  |  | 0.1 (0 to 1.2) | ↑ | ≤0.05 |
|  | Δ 28 Days* | Directional shift | Within P-value<sup>‡</sup> | Δ 28 Days* | Directional shift | Within P-value<sup>‡</sup> | Δ 28 Days* | Directional shift | Within P-value<sup>‡</sup> |
| *Enterocloster* | 0 (0 to 0.1) | ↑ | 0.07 |  |  |  |  |  |  |
| *Eubacterium_F* |  |  |  | 0.1 (0.1 to 0.3) | ↑ | 0.09 |  |  |  |
| *Eubacterium_J* |  |  |  |  |  |  | 0 (-0.2 to 0.1) | ↑ | 0.1 |
| *Faecalibacterium* | 3.2 (-0.8 to 6.2) | ↑ | 0.08 |  |  |  |  |  |  |
| *Fimenecus* |  |  |  | -0.1 (-0.6 to 0.2) | ↓ | 0.1 | -0.6 (-1.1 to -0.2) | ↓ | 0.06 |
| *Fusicatenibacter* |  |  |  |  |  |  | -0.6 (-0.8 to 0) | ↓ | 0.06 |
| *G11* |  |  |  | 0.3 (0.1 to 0.5) | ↑ | 0.06 |  |  |  |
| *Hominicoprocola* |  |  |  |  |  |  | 1.1 (0.2 to 1.9) | ↑ | 0.1 |
| *Howardella* |  |  |  | -0.1 (-0.1 to -0.1) | ↓ | ≤0.05 |  |  |  |
| *Intestinibacter* |  |  |  | 0 (-0.1 to 0) | ↓ | 0.06 |  |  |  |
| *Lachnoclostridium_B* |  |  |  |  |  |  | 0 (-0.2 to 0) | ↑ | 0.09 |
| *Lachnospira* | 0 (0 to 0.1) | ↑ | ≤0.05 | 0 (0 to 0.1) | ↑ | 0.1 |  |  |  |
| *Lawsonibacter* |  |  |  | 0 (0 to 0.1) | ↑ | 0.1 |  |  |  |
| *Limiplasma* |  |  |  | -0.1 (-0.1 to 0) | ↓ | ≤0.05 |  |  |  |
| *Limisoma* |  |  |  |  |  |  | 0.4 (0.1 to 0.5) | ↑ | 0.07 |
| *Megasphaera_A_38685* |  |  |  |  |  |  | 1.2 (1 to 3) | ↑ | 0.07 |
| *Mogibacterium_A* |  |  |  | -0.9 (-1.1 to -0.6) | ↓ | ≤0.05 |  |  |  |
| *Paraprevotella* | 0.1 (0.1 to 0.1) | ↑ | ≤0.05 | 0.2 (0.1 to 0.4) | ↑ | ≤0.05 |  |  |  |
| *Parasutterella_564865* |  |  |  | -0.1 (-0.1 to 0.2) | ↑ | 0.08 |  |  |  |
| *PeH17* |  |  |  |  |  |  | -0.1 (-0.1 to 0.2) | ↓ | 0.07 |
| *Porcincola* |  |  |  | 0 (0 to 0.1) | ↑ | ≤0.05 |  |  |  |
| *Prevotella* | 0.3 (-0.5 to 2.6) | ↑ | ≤0.05 |  |  |  | 0.3 (0 to 4.2) | ↑ | ≤0.05 |
| *Romboutsia_B* |  |  |  |  |  |  | 0 (-0.5 to 0.2) | ↑ | 0.05 |
| *Roseburia_C* |  |  |  |  |  |  | 0.2 (0 to 0.6) | ↑ | 0.08 |
| *Ruminococcus_B* | 0 (-0.1 to 0) | ↓ | ≤0.05 |  |  |  | -3.5 (-4.5 to -2.2) | ↓ | ≤0.05 |
| *Ruthenibacterium* | -0.1 (-0.1 to 0) | ↓ | ≤0.05 |  |  |  |  |  |  |
|  | Δ 28 Days* | Directional shift | Within *P*-value<sup>‡</sup> | Δ 28 Days* | Directional shift | Within *P*-value<sup>‡</sup> | Δ 28 Days* | Directional shift | Within *P*-value<sup>‡</sup> |
| *Schaedlerella* |  |  |  |  |  |  | 0 (-0.1 to 0) | ↑ | 0.1 |
| *Sutterella* | 0.1 (0.1 to 0.2) | ↑ | ≤0.05 | 0 (-0.1 to 0.5) | ↑ | ≤0.05 |  |  |  |
| *Terrisporobacter* |  |  |  |  |  |  | 0 (-0.1 to 0) | ↑ | ≤0.05 |
| *Turicibacter* |  |  |  | 0 (-0.1 to 0) | ↑ | 0.07 |  |  |  |
| *UBA6382* |  |  |  | 0 (0 to 0.2) | ↑ | 0.09 |  |  |  |
| *Wujia* |  |  |  | 0.1 (0 to 0.3) | ↑ | 0.09 |  |  |  |

\*Percent change in relative abundance for each taxon from baseline (Day 0) to Day 28. For each participant, the percentage increase or decrease was calculated as the difference between Day 28 and Day 0 values.
<sup>†</sup>Samples were collected from each participant at two time points: baseline (Day 0) and Day 28. The total participant analyzed for HD effera® were n=14, LD effera® were n=15, bLF were n=14.
<sup>‡</sup>Multivariate analysis by linear models (MaAsLin 3.0)<sup>38</sup> was used to determine associations between visit and the relative abundance of bacterial taxa within each treatment. The relative abundance data were log2-transformed; only the taxa with relative abundances ≥ 0.1% in more than 10% of the samples were included in the analyses. All *P*-values were corrected for multiple testing by the Benjamini-Hochberg false discovery rate (FDR). *P*-values ≤ 0.05 were considered statistically significant; 0.05 < *P* ≤ 0.10 were reported as trends toward significance.
Abbreviations: g/d, grams per day; ↑ and ↓, directional shifts in a genus at Day 28 compared to Day 0.

### 4. Volatile fatty acid concentrations

Bovine LF supplementation produced no significant within-group proportional changes in stool VFA composition (Table 4). On an absolute change basis, bLF led to significant or had near-significant median increases of total SCFA (53.9 mmol/kg), propionate (23.1 mmol/kg), BCFA (4.4 mmol/kg), isobutyrate (1.6 mmol/kg), and valerate (2.6 mmol/kg).

**Table 4.**
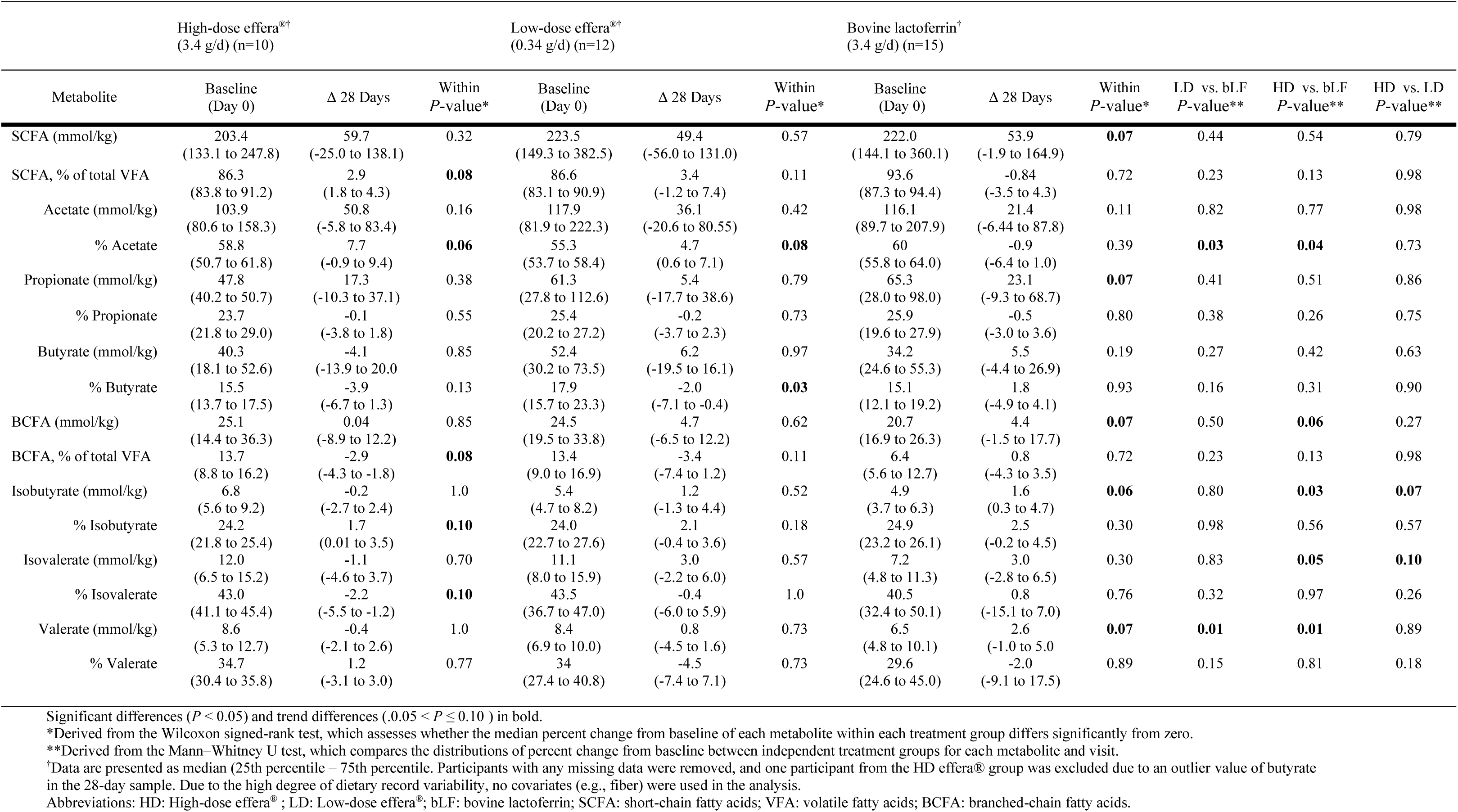
Effects of 28 days of lactoferrin supplementation on changes in proportional fecal volatile fatty acid concentrations of healthy adults.

In contrast to the bLF group, both effera^®^ groups produced no significant within-group absolute changes in stool VFA composition. Significant or near-significant proportional changes, however, were observed for numerous VFA in both effera^®^ groups (Table 4). The proportion of total SCFAs (of total VFA) also had a trend increase in the HD effera^®^ group (*P* < 0.10) (Supplementary Figure A3). In the HD and LD effera^®^ groups, fecal acetate increased by 7.7% and 4.7%, respectively, relative to baseline (*P* < 0.10). Proportionally, fecal butyrate in the LD effera^®^ group decreased by 2.0% (*P* < 0.05); however, no absolute butyrate changes were observed (Supplementary Figure A4). The HD effera^®^ group showed trend proportional increases in overall BCFA and isobutyrate. Isovalerate, on the other hand, showed a proportional decrease.

Between-group analysis showed that the proportional increase in stool acetate was greater in both effera^®^ groups than in the bLF group (*P* < 0.05 for HD effera^®^ vs bLF; *P* < 0.05 for HD effera^®^ vs bLF). In addition, between-group differences were observed in the absolute total BCFA and individual BCFA including valerate, isovalerate, and isobutyrate between bLF and HD effera^®^. A significant difference between the LD effera^®^ and bLF group was found for valerate (Table 4).

The trend of increasing acetate after the 28-day treatment appeared to shift concentrations back towards baseline at both follow-up visits at Day 56 and Day 84 (Supplementary Figure A5). The increased acetate production in the LD and HD effera^®^ was not dose-responsive; however, among females, acetate was more consistently elevated after the 28-day supplementation compared to males (Supplementary Figure A6). We did not observe similar changes among males and females in either butyrate or propionate concentrations (Supplementary Figure A6).

## 4. Discussion

In this exploratory microbiome study using fecal samples collected from generally healthy adults consuming free-living diets, supplementation with effera^®^ or bLF for four weeks did not induce large-scale restructuring of the gut microbiome, as assessed by alpha and beta-diversity metrics.

Alpha-diversity analyses showed no significant changes in richness, evenness, or phylogenetic diversity over time and no differences between bLF or HD effera^®^ and LD effera^®^. The stability of alpha-diversity observed in a healthy adult population mirrors findings from other human intervention trials involving LF. In a study of healthy elderly women supplemented with 1 g/day of bLF for three weeks, Konstanti et al., reported no significant differences in alpha-diversity (utilizing Faith’s PD and Shannon index).^44^ Similarly, a randomized trial in toddlers (12–18 months old) receiving 1 g/day of bLF for six months found no significant differences in alpha-diversity metrics between the treatment and placebo groups.^19^ These data suggest that among participants with a relatively stable or established microbiome, LF supplementation preserves community richness and evenness. From a safety and tolerability standpoint, the absence of marked reductions in alpha-diversity reemphasizes LF as a safe dietary protein that maintains microbiome diversity metrics over a 28-day dietary intervention.

Analysis of beta-diversity revealed that supplementation with effera^®^ maintains the gut microbial community in healthy adults. PERMANOVA results indicated no significant main effect of treatment or treatment-by-visit interaction for either weighted or unweighted UniFrac distances, suggesting that neither LD effera^®^ nor HD effera^®^ induced disruptions to the gut microbiota. These findings align with previous investigations in healthy populations, such as the study by Konstanti et al., which found no significant differences in beta-diversity between healthy elderly women supplemented with either bLF or a placebo.^44^ Similarly, a trial with toddlers supplemented with bLF found no significant clustering by treatment using Aitchison distance.^19^ In preterm infants, targeted 16S analyses have reported that bLF supplementation does not overtly disrupt microbiota development supporting a “selective modulation without destabilization” model.^45^ Together, these infant data support the idea that LF might act as a context-dependent modulator, most evidently at the level of specific taxa and functions, rather than as the primary driver of widespread community remodeling, which aligns with the overall stability observed here in adults.^21^

Against this backdrop of overall stability, an effect of visit was observed, indicating temporal shifts, and stratification by treatment revealed distinct patterns of stability. Supplementation with bLF was associated with significant visit-related shifts in beta diversity, as measured by weighted UniFrac distances. Weighted UniFrac accounts for the relative abundance of taxa, suggesting that bLF may have induced shifts in the proportions of specific microbial populations over the 28-day treatment period. This is consistent with findings of Chichlowski et al., who reported subtle but significant differences in beta-diversity (Jaccard distance) among infants fed bLF-enriched formula, driven by changes in low-abundance species.^46^ Similarly, Embleton et al., reported small but significant differences in community composition between bLF and placebo groups in preterm infants.^18^

At the phylum level, the general shift towards a higher relative abundance of *Bacteroidota* is important for host health and aligns with the previously reported modulatory effects of LF on intestinal microbiota.^15,47–49^ Moreover, this increase in *Bacteroidota* corresponded with a significant decrease of *Bacillota_I* in the LD effera^®^ groups and a non-significant decrease in HD effera^®^. Although the changes did not always reach significance, shifts in these major microbiome phyla have been observed in previous animal and human studies.^8,48,50^

Despite a broad relative abundance decrease in the *Bacillota* phyla, effera^®^ induced increases in several butyrate-producing genera, which support a healthy gut environment. Both effera^®^ doses increased the abundance of *Lachnospira*, a key genus known for fermenting dietary fiber into butyrate and acetate.^51^ The positive correlation of effera^®^ with *Lachnospira* may be related to the ability of effera^®^ to bind iron, as *Lachnospira*’s abundance is associated with low iron intake.^52^ Overall, *Lachnospira* contributes to gut health by producing fermentation metabolites (butyrate/acetate) that nourish colonocytes and support the intestinal barrier. ^53^ These observations are notable in light of longitudinal infant cohorts in which early-life depletion of specific fermentative lineages, including *Lachnospira*, has been associated with increased later risk of wheeze/asthma, implicating early functional capacity as a potential determinant of immune trajectories.^53^However, infant microbiome assembly is constrained by human-milk glycans, rapid ecological succession, and strong clinical confounding attributes, such that adult-directionality cannot be assumed to translate across life stages. Accordingly, infant-focused translation of LF should prioritize integrated endpoints that couple composition with function (absolute SCFAs, lactate, pH, and cross-feeding signatures) and, where feasible, host readouts of barrier and immune status.^54^

*Paraprevotella* also rose significantly in both effera^®^ groups. *Paraprevotella* ferments non-digestible complex carbohydrates and produces SCFAs (notably succinate and acetate).^55^ Higher abundance of *Paraprevotella* has been associated with adherence to fiber-rich diets.^56^ Consistent increases in *Paraprevotella* across both the effera^®^ groups signals a shift favoring *Bacteroidetes* (as was corroborated in the phylum data). These changes support the idea that LF supplementation can modulate the microbiome in a prebiotic-like manner.

Similarly, the LD effera^®^ increased the genus *Eubacterium* which is known to produce butyrate and acetate and promote colonocyte health and dampen inflammation.^52^ *Eubacterium* helps maintain a eubiotic state and has been associated with protection against conditions inflammatory bowel disease and insulin resistance.^57,58^ The enhancement of SCFA producers such as *Lachnospira* and *Eubacterium* by effera^®^ could thus have positive implications, potentially contributing to improved gut barrier integrity and immune balance.

*Faecalibacterium* abundance showed a trend increase in HD effera^®^ and a near trend increase in the LD effera^®^ after 28 days of treatment. This major butyrate producer is well known for supporting a healthy inflammatory response^59–61^ and enhancing the intestinal epithelial barrier and maintaining gut homeostasis.^62^ This finding is encouraging as effera^®^ supported the increase in the relative abundance of a genera that contains *F. prausnitzii*, a next-generation probiotic species.^60^

We report the HD effera^®^ group was significantly associated with an increase in *Agathobacter*, which includes the species *A. rectalis*. *A. rectalis* is one of the most abundant butyrate-producing commensals in healthy adults and has recently been shown to alleviate intestinal and even neurological inflammation in experimental settings through its production of butyrate.^63^ An effera^®^-driven rise in *Agathobacter* is again consistent with enriching for beneficial fiber fermenters. HD effera^®^ also significantly increased *Alistipes*. This finding was in agreement with a study that showed how different degrees of LF iron saturation supported the growth of commensals.^8^ Another study in a preclinical obese mouse model showed bLF increased *Alistipes*, which was associated with a healthy inflammatory response in obese mice.^48^

Interestingly, we found that both bLF and effera^®^ (particularly LD effera^®^) consistently suppressed several genera in the family *Lachnospiraceae*, including *Dorea* and *Blautia*, which are normal commensals in a balanced gut ecosystem.^64,65^ However, an overabundance of *Dorea* has been linked to gut dysbiosis and adverse host phenotypes, including type 2 diabetes.^66^ *Blautia*, on the other hand, is often considered beneficial. It is a major butyrate producer in the gut of healthy individuals, and has been inversely associated with visceral fat accumulation and metabolic disease risk.^64^ The mechanism by which *Blautia* may have been reduced is through iron sequestration by LF. *Blautia* is known to prefer iron-rich growth environments,^67,68^ which may have been impeded by LF’s ability to sequester iron.

HD effera^®^ and bLF were significantly associated with *Prevotella*. *Prevotella* is known to be associated with plant-based diets and has the potential for protection against food allergy development when observed in the pregnant woman.^69^ We found that bLF, while sharing some microbiome-modulation effects with effera^®^, had distinct impacts on the healthy adult gut microbiome. bLF caused a significant increase of a major butyrate producer *Roseburia*. In a piglet model, Hu et al. showed significant increases of *Roseburia*.^15^

BLF significantly increased beneficial genera including *Bacteroides, Cryptobacteroides*, and *Eisenbergiella. Eisenbergiella* is a new genus but it has been identified as a butyrate producer that may support gut barrier function and support a healthy inflammatory response.^70^ Overall, the genus *Bacteroides* are commensal organisms, however there are scenarios where they act as opportunistic pathogens in specific scenarios where the immune system is compromised.^71^

On the metabolite side, a trend toward increased acetate proportion after 28-day treatment with HD and LD effera^®^ supplementation and a significant increase, compared to bLF, suggest a potentially meaningful shift in microbial metabolic activity. Acetate is the most abundant SCFA in the colon,^72^ constituting more than 50% of the total SCFAs in our study. Acetate is found in high amounts in infants fed human milk compared to those fed infant formula, due to the higher abundance of Bifidobacteria strains in human milk-fed infants.^73–75^ The trend in acetate levels appeared consistent with our reported multiple genus shifts rather than a single genus. The directional change was consistent with the capacity of hLF to exert an effect on the human gut microbiota, which is of interest given that acetate is the predominant SCFA in human milk-fed infants.^75^

It is also noteworthy that in our exploratory study, sex-linked acetate production trend was mainly found among female participants, who appeared to respond to HD and LD effera^®^ with greater increases in acetate, whereas men showed smaller or no changes. While our sample size was limited, sex hormones and other physiological differences between females and males are known to shape the gut ecosystem across the lifespan.^76^ Further studies on controlled dietary conditions could further resolve these observations on both effera^®^ doses compared to the bLF cohort. Although this study was conducted in healthy adults, the distinct metabolic profiles observed between the LF groups—where bLF drove absolute increases in total SCFA and BCFAs while effera^®^ drove proportional shifts toward acetate—mirror the metabolic divergence observed between formula-fed and human milk-fed infants.^77^ From a translational perspective, acetate is a compelling functional endpoint because it typically dominates fecal SCFAs in milk-fed infancy, and acetate-enriched profiles are repeatedly associated with breastfeeding-associated fermentation networks.^81^ Future infant studies with effera^®^ would therefore benefit from pairing taxonomic profiling with quantitative metabolomics (absolute SCFAs, lactate, succinate, where feasible) and linking these to barrier/immune readouts to determine whether LF preferentially tunes microbial function rather than global community structure.

The significant absolute increases in total BCFAs and two individual BCFAs (i.e., isobutyrate and valerate) in the bLF group act as a biomarker of proteolytic fermentation, a process driven by the microbial catabolism of branched-chain amino acids such as valine, leucine, and isoleucine.^78^ In contrast, the effera^®^ groups exhibited a metabolic profile analogous to the human milk-fed infant, defined by significantly higher relative proportion of acetate. This distinction may be physiologically relevant because, while carbohydrate fermentation is generally beneficial, the fermentation of proteins yields a more diverse range of metabolites, including ammonia, BCFAs, and phenolic compounds.^79^ Some of these compounds can be deleterious to the colonic epithelium at high concentrations.^79,80^

Taken together, these compositional and metabolite-level observations highlight both the overlapping as well as distinct effects of hLF produced by *K. phaffii* and bLF on the human gut microbiome and fecal VFA. Some similarities are probably due to the evolutionary conservation of LF across species.^82,83^ However, species-specific structural features probably explain the observed differences. In the original study, on which this primary analysis is based, Peterson et al. showed that bLF induced anti-bLF antibodies in numerous participants, whereas both doses of effera^®^ did not show any increase in antibodies at any of the post-treatment follow-up visits.^27^ These observations suggest that, despite 69% amino acid sequence homology, the immune system can distinguish between bLF and effera^®^. Moreover, it is known that the bioactive peptides derived from hLF and bLF differ slightly in amino acid composition, charge, and hydrophobicity, resulting in distinct antimicrobial activities.^84^ Finally, similarities and differences in iron saturation and glycosylation profiles between effera^®^ and bLF may also contribute to the observed overlapping and divergent microbial shifts.^8,83^

In conclusion, this hypothesis-generating study provides evidence that LD and HD effera^®^ maintained a diverse microbiome as measured by alpha- and beta-diversity, promoted proportional increases in SCFA production (particularly acetate), and enriched numerous potentially beneficial genera in healthy adults. Future adult clinical studies should look at other larger participant cohorts (larger “n” and controlled diet) to corroborate these reported findings on taxa shifts and VFA profiles. In addition, investigating effera^®^ hLF in infant populations may further clarify its potential to help close the compositional gap between formula-fed and breastfed infants, at both metabolite and microbial levels.

## Supplementary Materials

### Author Contributions

Author Contributions: RDP: Conceptualization, visualization, resources, writing- original draft, supervision, project administration; JVM: Methodology, formal analysis, writing- review and editing; NK: Conceptualization; SMD: Conceptualization, writing- review and editing, supervision, project administration; funding acquisition; MW: Conceptualization, formal analysis; writing- review and editing, RND: Methodology, formal analysis; AJC: Conceptualization, resources, supervision.

### Funding Information

The authors disclosed receipt of the following financial support for the research, authorship, and publication of this article: Helaina Inc. funded this research and was involved in the study design, interpretation of data, writing of the report, and the decision to submit the report for publication.

### Institutional Review Board Statement

Ethics approval statement: These studies were approved by the WIRB-Copernicus Group Institutional Review Board (protocol: MB-2305, 8/16/2023).

### Informed Consent Statement

Written informed consent was obtained from each participant prior to testing.

### Data Availability Statement

The datasets generated and analyzed during the current study are not publicly available due to proprietary and confidential business considerations of Helaina Inc. Requests for access to the data may be directed to the corresponding author and will be considered on a case-by-case basis, subject to applicable confidentiality agreements.

## Acknowledgments

Medical writing and editing support were provided by Carolyn Alish, of Sterling Medical Communications LLC. VFA analyses were conducted by Laura Bauer at the University of Illinois at Urbana-Champaign.

## Conflicts of Interest and Other Ethics Statements

Julian van der Made, Nicole Kaplan, Ross Peterson, and Anthony Clark are employees of Helaina Inc. and receive equity or stocks in the company. Sharon Donovan and Ryan Dilger received funding from Helaina Inc. to perform the microbiome and VFA analyses, respectively.

