## Supplemental Material for "Effects of Human Lactoferrin (effera^®^) at Two Doses versus Bovine Lactoferrin on the Adult Gut Microbiome and Fecal Short-Chain Fatty Acids: A Randomized, Double-Blind Trial"

### Supplementary Figures

**Supplementary Table A1.** Alpha-diversity measures (observed features, Shannon entropy, evenness, and Faith's phylogenetic diversity) across treatment groups, from baseline (Day 0) to Day 28.

| | Baseline (Day 0) | | | Post Treatment (Day 28) | | | *P* value | | |
|---|---|---|---|---|---|---|---|---|---|
| | HD efferà® (n=14) | LD efferà® (n=15) | bLF (n=14) | HD efferà® (n=14) | LD efferà® (n=15) | bLF (n=14) | Treatment | Visit | Interaction |
| Observed features | 662.9 ± 53.1 | 793.7 ± 46.7 | 699.3 ± 61.7 | 705.6 ± 69.4 | 793.5 ± 62.5 | 712.4 ± 58.0 | 0.3453 | 0.3356 | 0.6452 |
| Shannon | 7.26 ± 0.21 | 7.78 ± 0.10 | 7.45 ± 0.16 | 7.35 ± 0.22 | 7.63 ± 0.18 | 7.56 ± 0.17 | 0.2008 | 0.8581 | 0.4629 |
| Evenness | 0.78 ± 0.01 | 0.81 ± 0.01 | 0.80 ± 0.01 | 0.78 ± 0.01 | 0.80 ± 0.01 | 0.80 ± 0.01 | 0.1956 | 0.9389 | 0.4439 |
| Faith's PD | 23.94 ± 1.51 | 27.16 ± 1.56 | 24.03 ± 2.03 | 25.17 ± 1.69 | 27.06 ± 1.57 | 24.74 ± 1.79 | 0.3908 | 0.2548 | 0.5876 |

Values are expressed as Mean ± SEMs;
Data were analyzed using lmer function of lme4 R package. The statistical model included treatment, visit and the interaction between treatment and visit. Total fiber intake (mean of 6 days for each participant) was included in the model as a covariate.
Abbreviations: bLF: bovine lactoferrin; HD: High-dose efferà® (3.4 g/d); LD: Low-dose efferà® (0.34 g/d); PD, phylogenetic diversity.

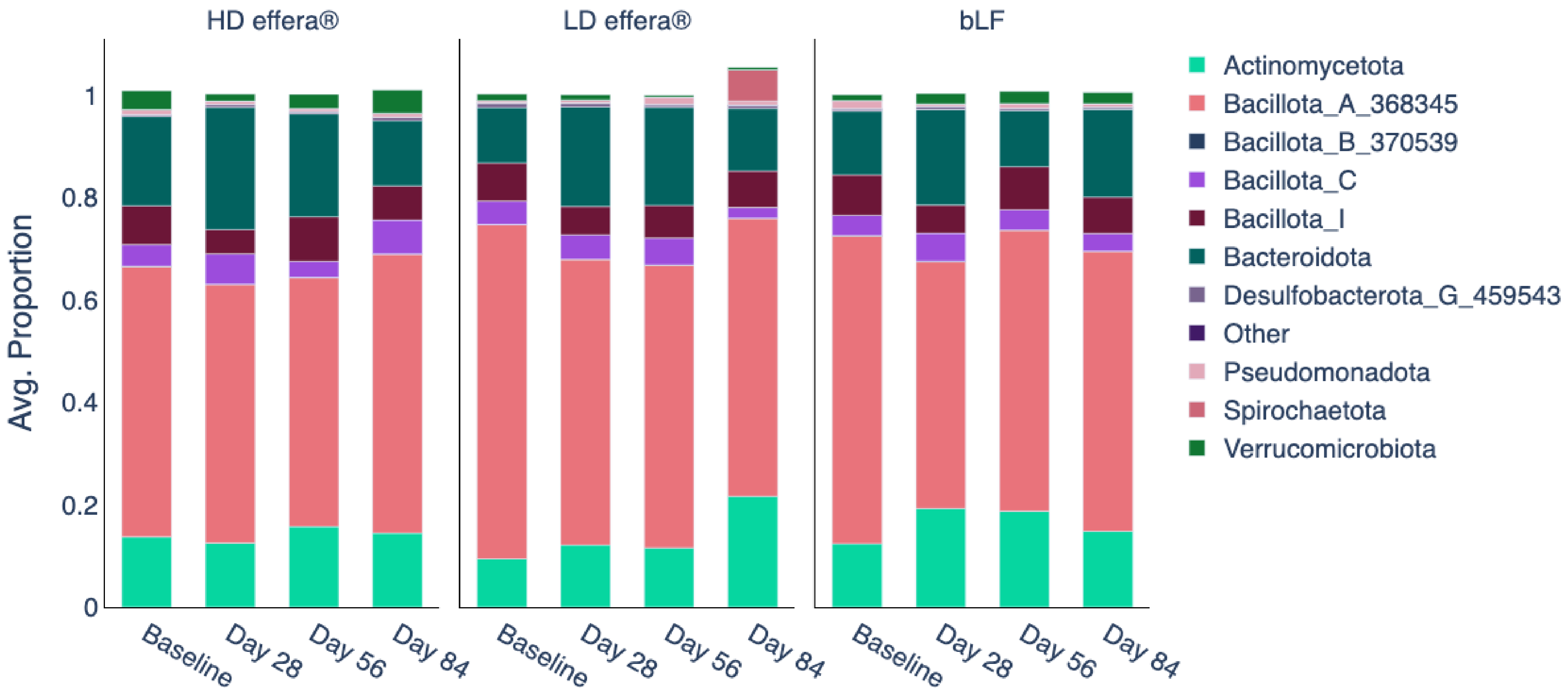


**Supplementary Figure A1.** Taxonomic stacked bar plots of mean abundance of phyla at baseline (Day 0) through supplementation (Day 28) and washout periods (Day 56 and Day 84).
Counts were normalized to their library size; the 10 most abundant phyla were plotted; "Other" phyla included all remaining phyla not in the top 10. Average proportions were individual relative abundances, averaged across the treatment group. Abbreviations: HD, high-dose effera®; LD, low-dose effera®; bLF, bovine lactoferrin.
The total participants analyzed for HD effera® (3.4 g/d) were n=14, LD effera® (0.34 g/d) were n=16, bLF (3.4 g/d) were n=15.

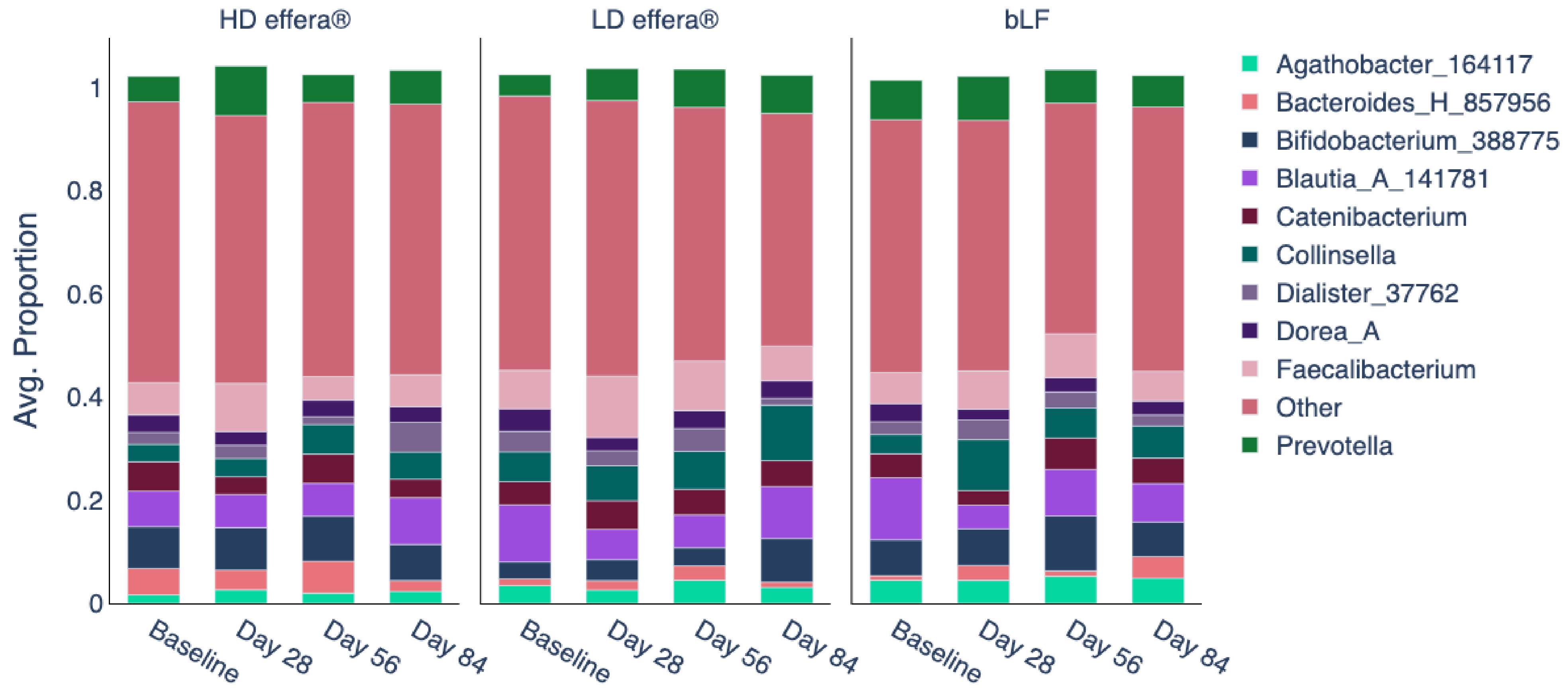


**Supplementary Figure A2.** Taxonomic stacked bar plots of mean abundance of genera at baseline (Day 0) through supplementation (Day 28) and washout periods (Day 56 and Day 84).
Counts were normalized to their library size; the 10 most abundant genera were plotted; "Other" genera included all remaining genera not in the top 10. Average proportions were individual relative abundances, averaged across the treatment group.
Abbreviations: HD, high-dose effera®; LD, low-dose effera®; bLF, bovine lactoferrin.
The total participants analyzed for HD effera® (3.4 g/d) were n=14, LD effera® (0.34 g/d) were n=16, bLF (3.4 g/d) were n=15.

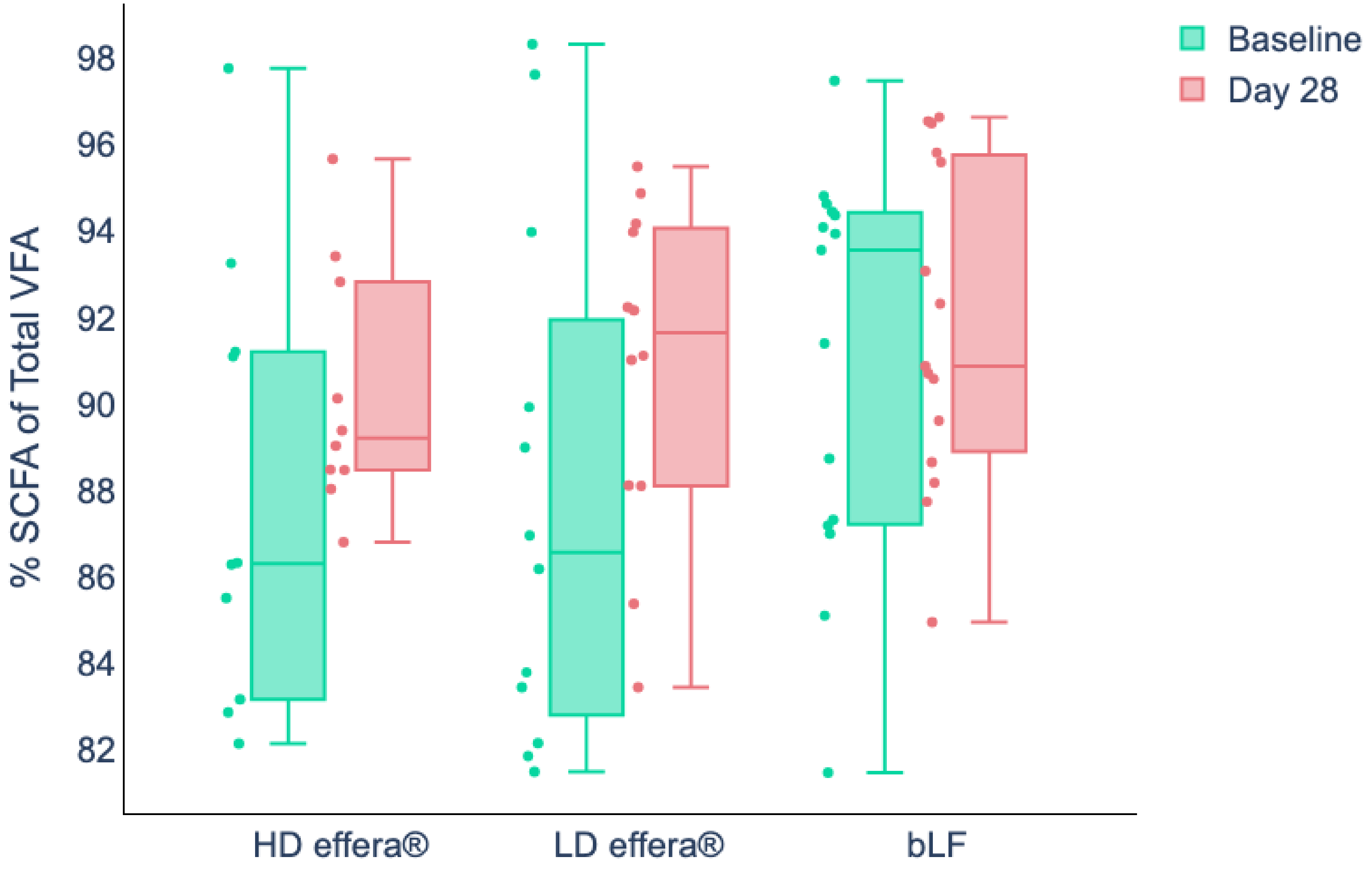


**Supplementary Figure A3.** Proportion of SCFAs within the total fecal fatty acid content at baseline (Day 0 and Day 28) for each treatment group.
Data are normalized as the total SCFA of the total VFA.
High-dose effera® (3.4 g/d) (n = 10); Low-dose effera® (0.34 g/d) (n = 12); Bovine Lactoferrin (3.4 g/d) (n = 15).
Abbreviations: SCFA, short-chain fatty acids; VFA, volatile fatty acids; HD, high-dose effera®; LD, low-dose effera®; bLF, bovine lactoferrin.

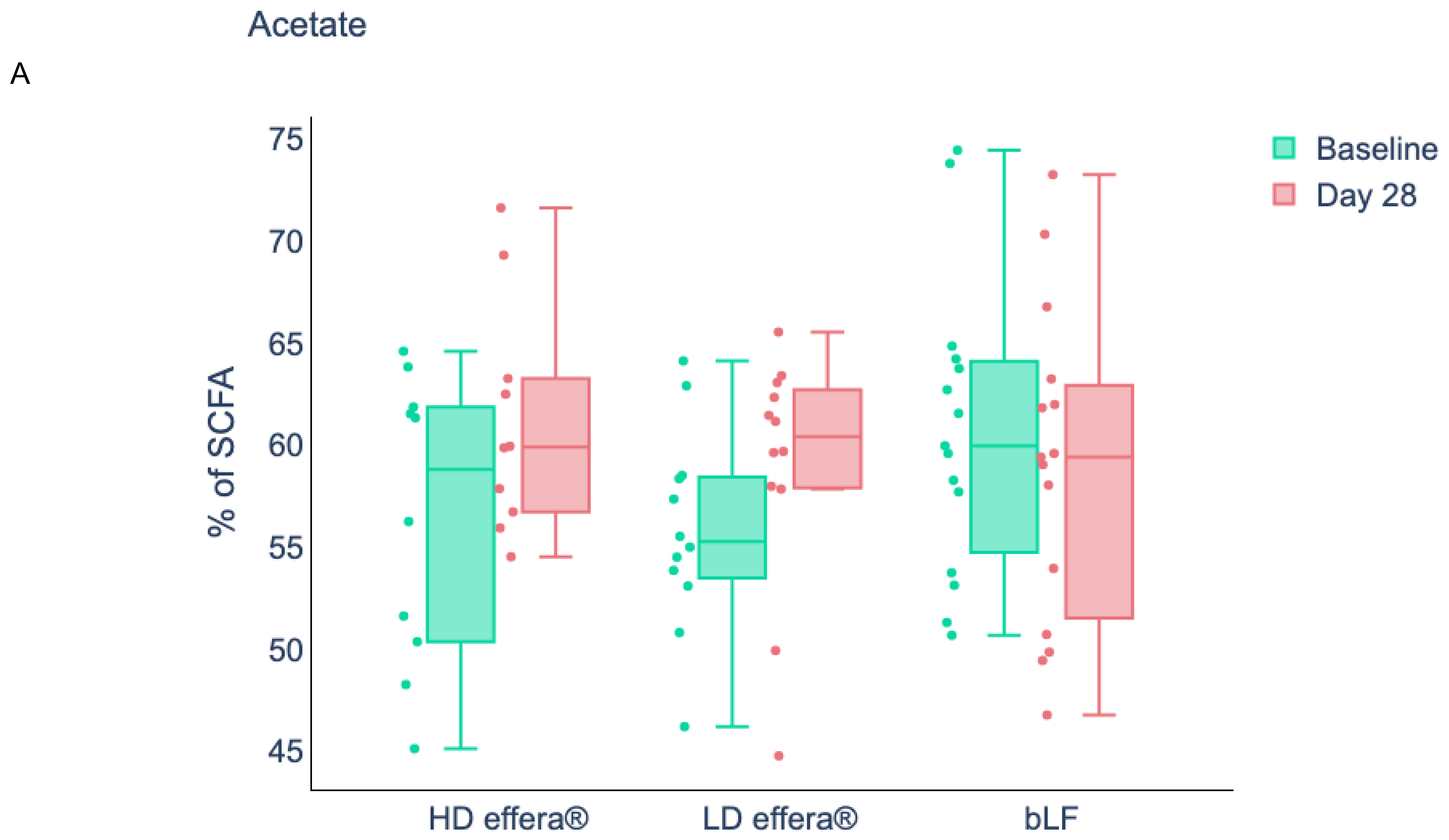


**Supplementary Figure A4.** Proportion of SCFA as A) acetate, B) propionate, and C) butyrate at baseline (Day 0 and Day 28) for each treatment group.
High-dose effera® (3.4 g/d) (n = 10); Low-dose effera® (0.34 g/d) (n = 12); Bovine Lactoferrin (3.4 g/d) (n = 15).
Abbreviations: HD, high-dose effera®; LD, low-dose effera®; bLF, bovine lactoferrin.

B

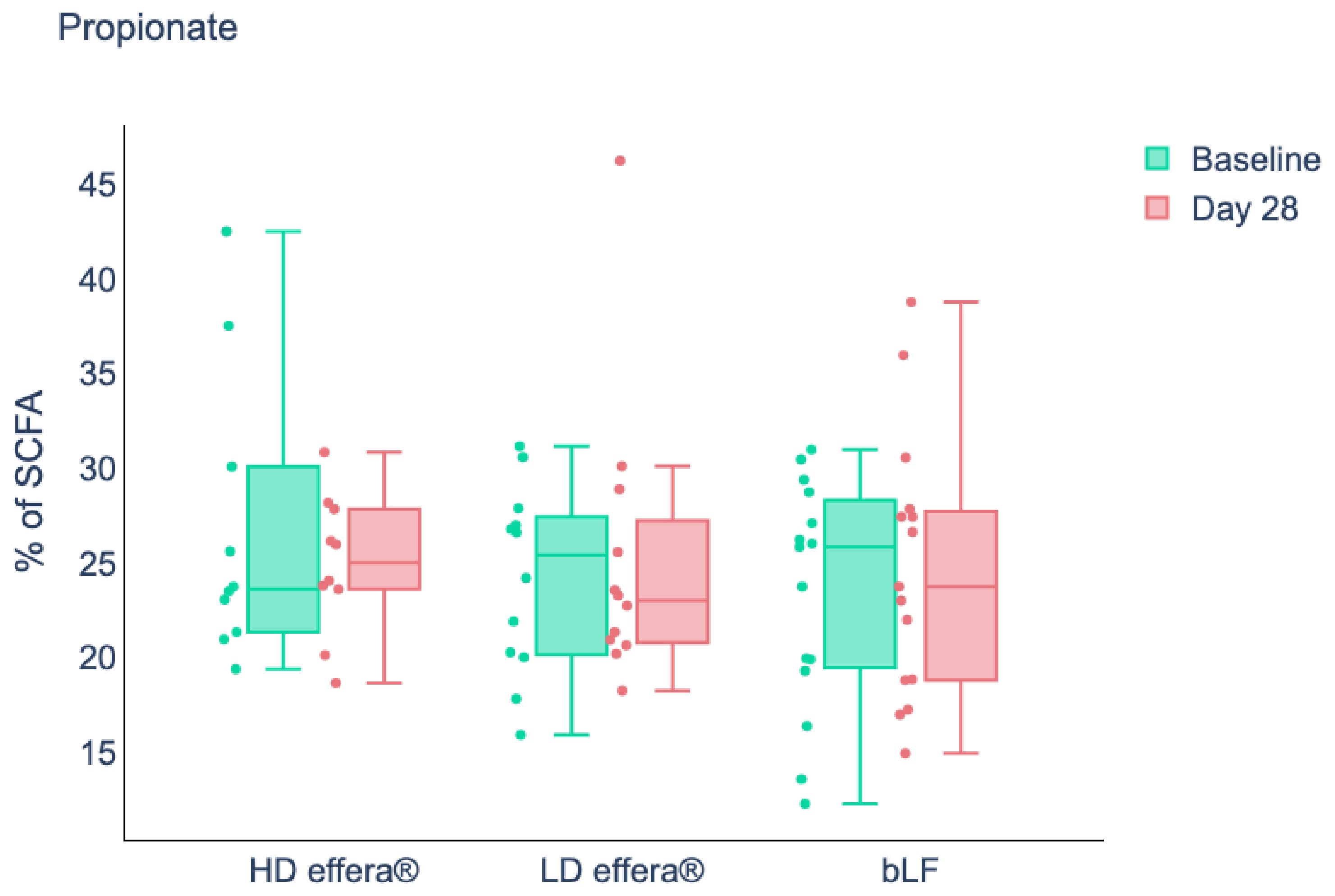


**Supplementary Figure A4.** Proportion of SCFA as A) acetate, B) propionate, and C) butyrate at baseline (Day 0 and Day 28) for each treatment group.
High-dose effera® (3.4 g/d) (n = 10); Low-dose effera® (0.34 g/d) (n = 12); Bovine Lactoferrin (3.4 g/d) (n = 15).
Abbreviations: HD, high-dose effera®; LD, low-dose effera®; bLF, bovine lactoferrin.

C

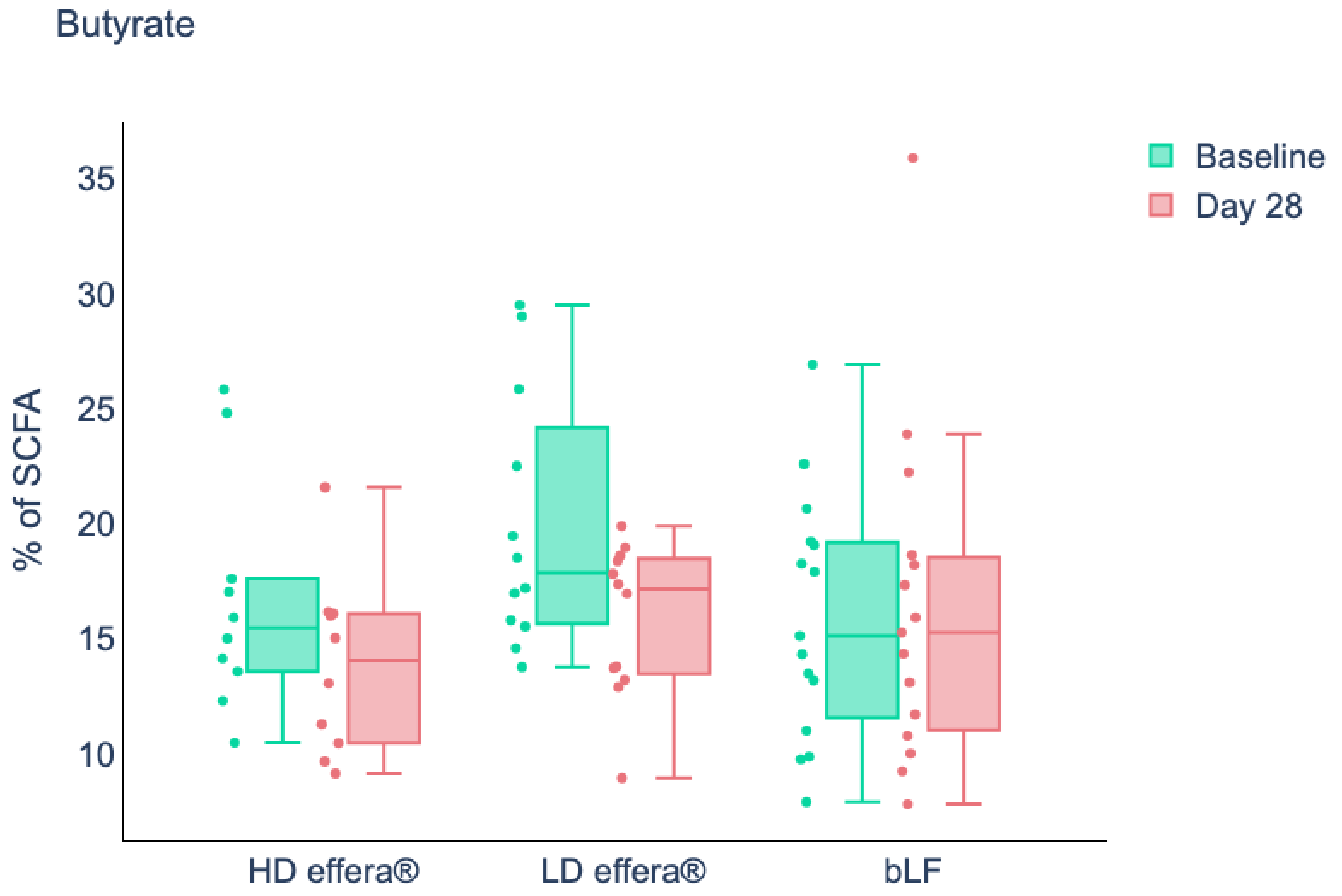


**Supplementary Figure A4.** Proportion of SCFA as A) acetate, B) propionate, and C) butyrate at baseline (Day 0 and Day 28) for each treatment group.
High-dose effera® (3.4 g/d) (n = 10); Low-dose effera® (0.34 g/d) (n = 12); Bovine Lactoferrin (3.4 g/d) (n = 15).
Abbreviations: HD, high-dose effera®; LD, low-dose effera®; bLF, bovine lactoferrin.

A

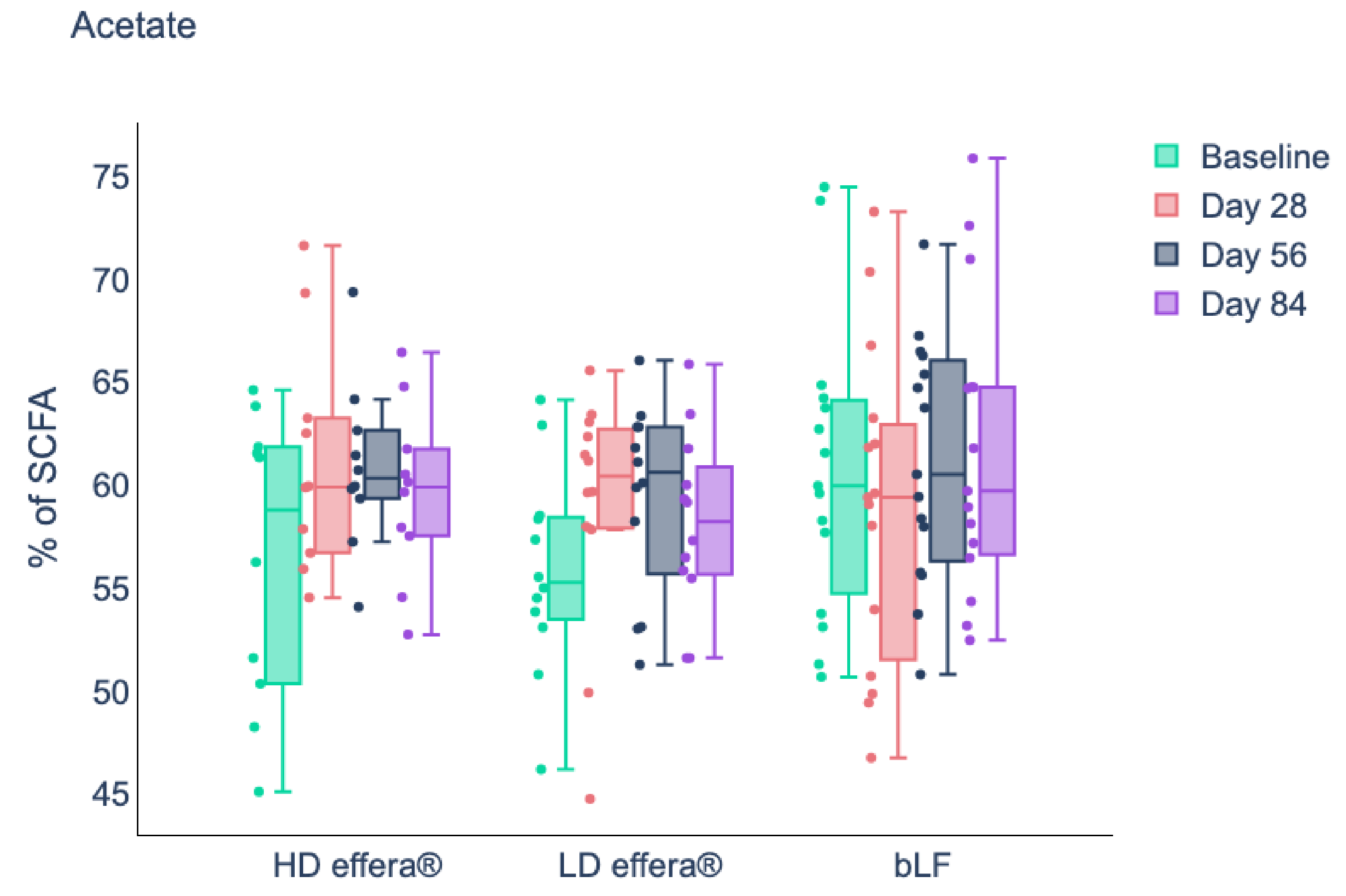


**Supplementary Figure A5.** Proportions of A) acetate, B) propionate, C) and butyrate at baseline (Day 0, Day 28, Day 56 and Day 84) for each treatment group. Abbreviations: HD, high-dose effera®; LD, low-dose effera®; bLF, bovine lactoferrin.

B

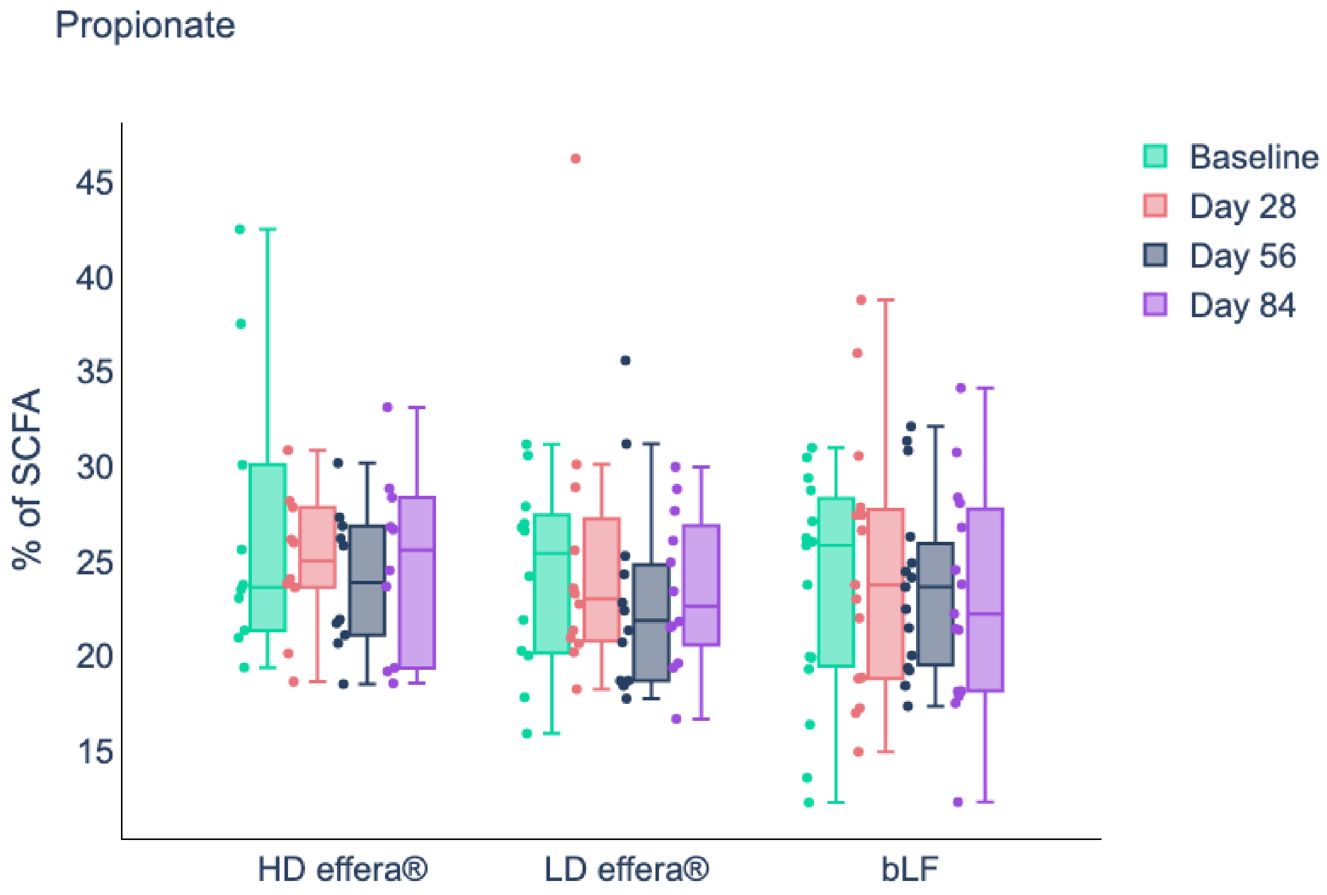


**Supplementary Figure A5.** Proportions of A) acetate, B) propionate, C) and butyrate at baseline (Day 0, Day 28, Day 56 and Day 84) for each treatment group. High-dose effera® (3.4 g/d) (n = 10); Low-dose effera® (0.34 g/d) (n = 12); Bovine Lactoferrin (3.4 g/d) (n = 15). Abbreviations: HD, high-dose effera®; LD, low-dose effera®; bLF, bovine lactoferrin.

C

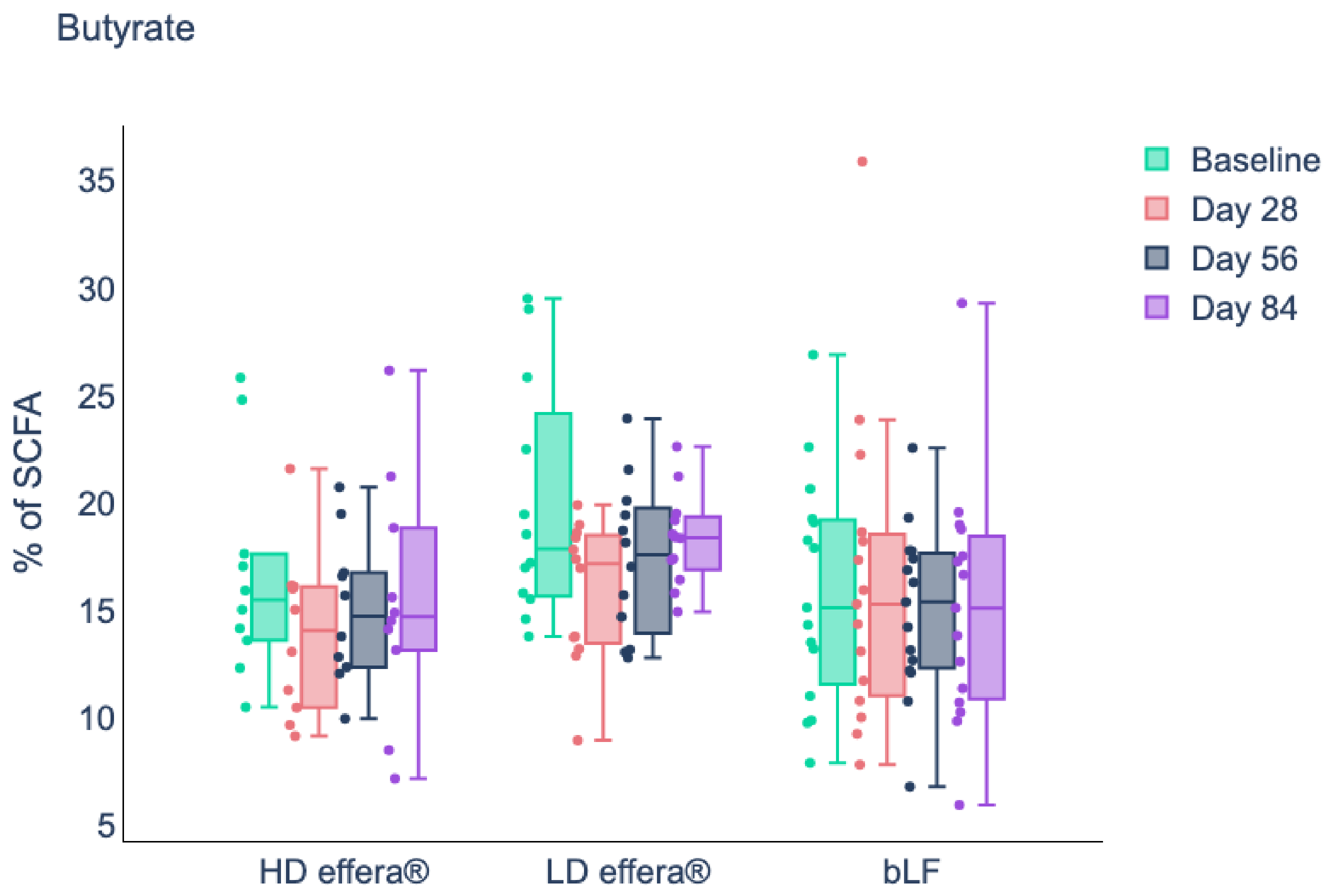


**Supplementary Figure A5.** Proportions of A) acetate, B) propionate, C) and butyrate at baseline (Day 0, Day 28, Day 56 and Day 84) for each treatment group. High-dose effera® (3.4 g/d) (n = 10); Low-dose effera® (0.34 g/d) (n = 12); Bovine Lactoferrin (3.4 g/d) (n = 15). Abbreviations: HD, high-dose effera®; LD, low-dose effera®; bLF, bovine lactoferrin.

A

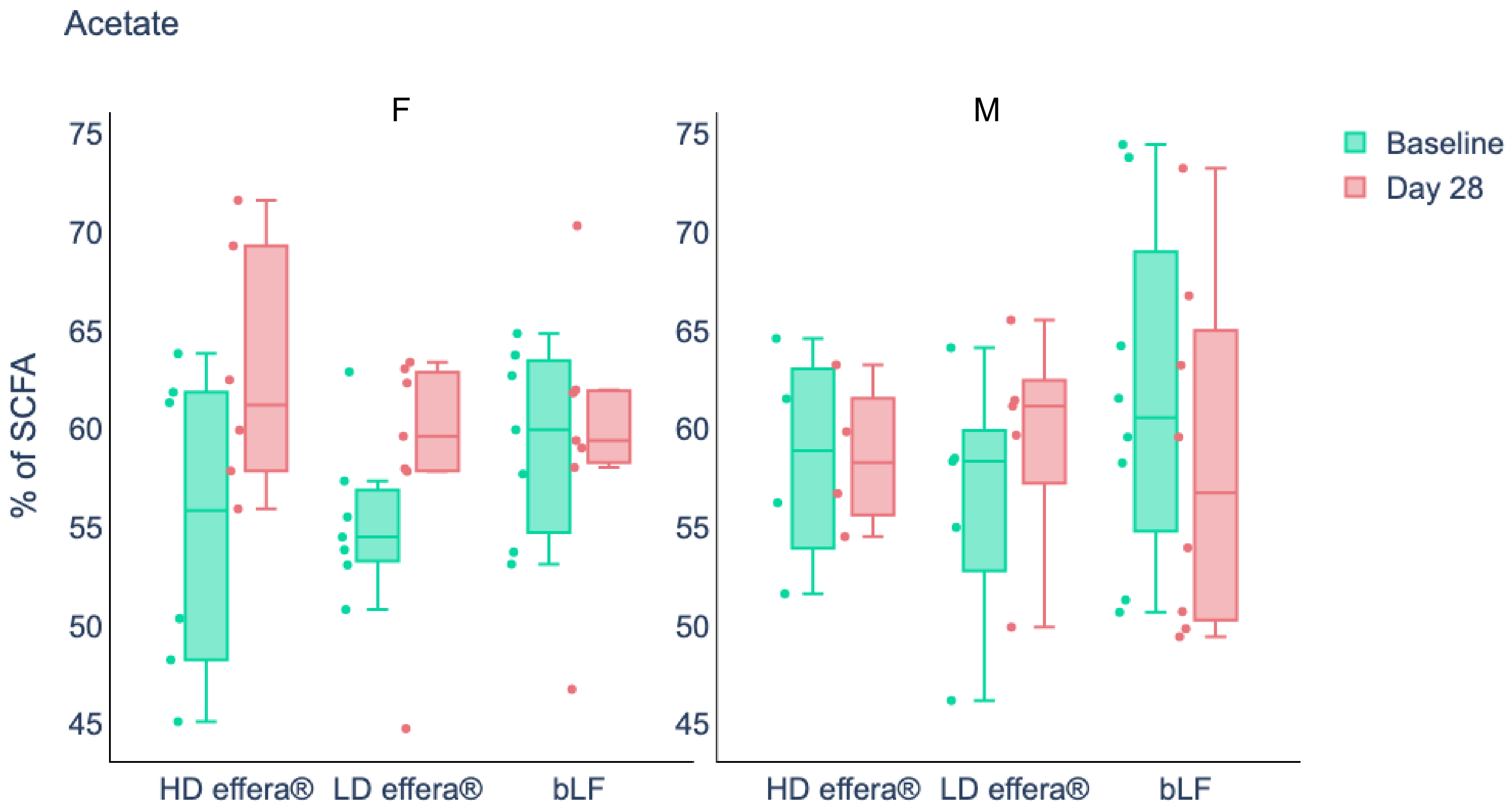


**Supplementary Figure A6.** Proportions of A) acetate, B) propionate, and C) butyrate at baseline (Day 0 and Day 28) for each treatment group by sex. High-dose effera® (3.4 g/d) (n = 10; ; F:6 M:4); Low-dose effera® (0.34 g/d) (n = 12; F:7 M:5); Bovine Lactoferrin (3.4 g/d) (n = 15; F:7 M:8). Abbreviations: HD, high-dose effera®; LD, low-dose effera®; bLF, bovine lactoferrin; F, female; M, male.

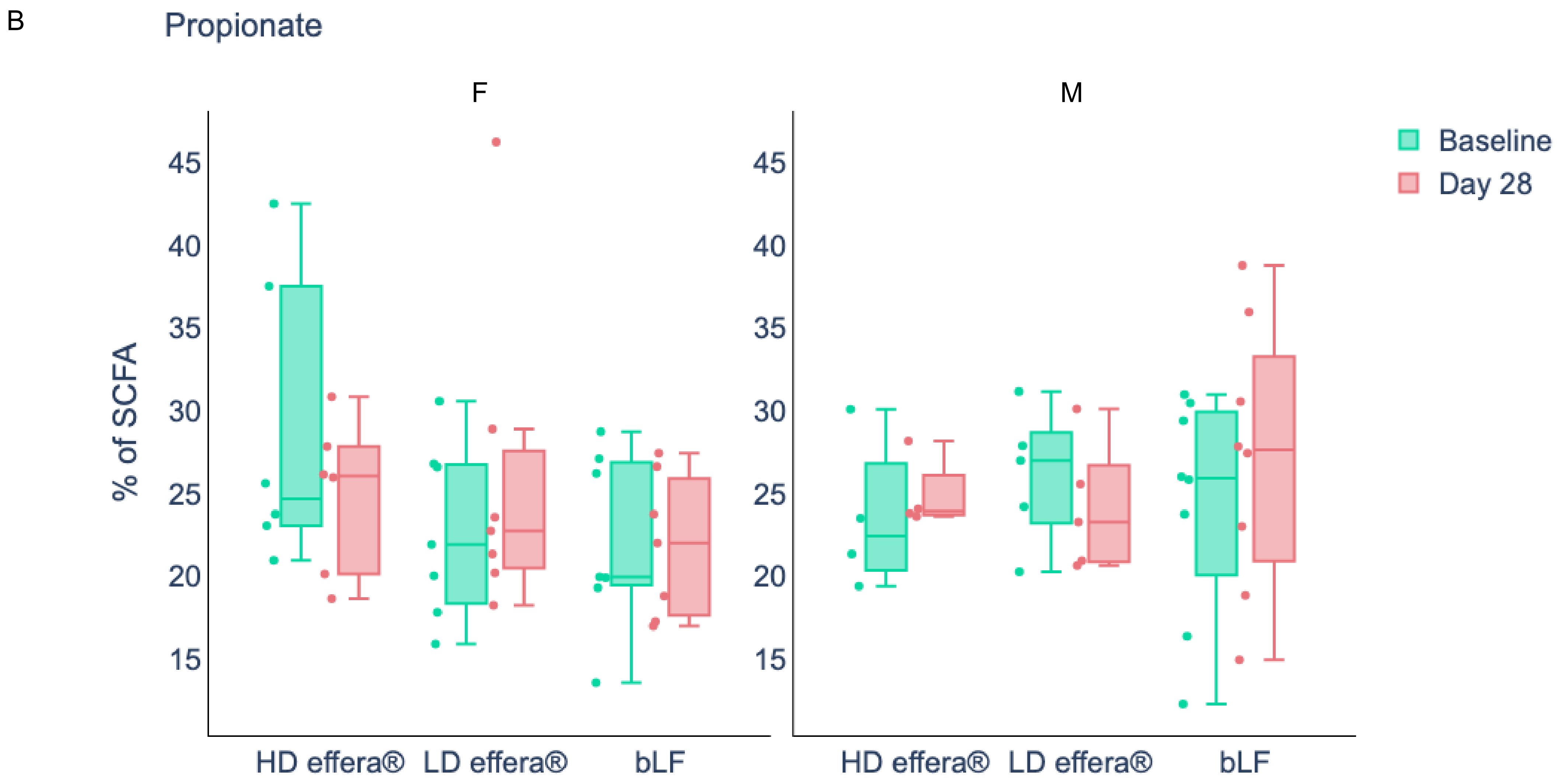


**Supplementary Figure A6.** Proportions of A) acetate, B) propionate, and C) butyrate at baseline (Day 0 and Day 28) for each treatment group by sex. High-dose effera® (3.4 g/d) (n = 10; ; F:6 M:4); Low-dose effera® (0.34 g/d) (n = 12; F:7 M:5); Bovine Lactoferrin (3.4 g/d) (n = 15; F:7 M:8). Abbreviations: HD, high-dose effera®; LD, low-dose effera®; bLF, bovine lactoferrin; F, female; M, male.

C

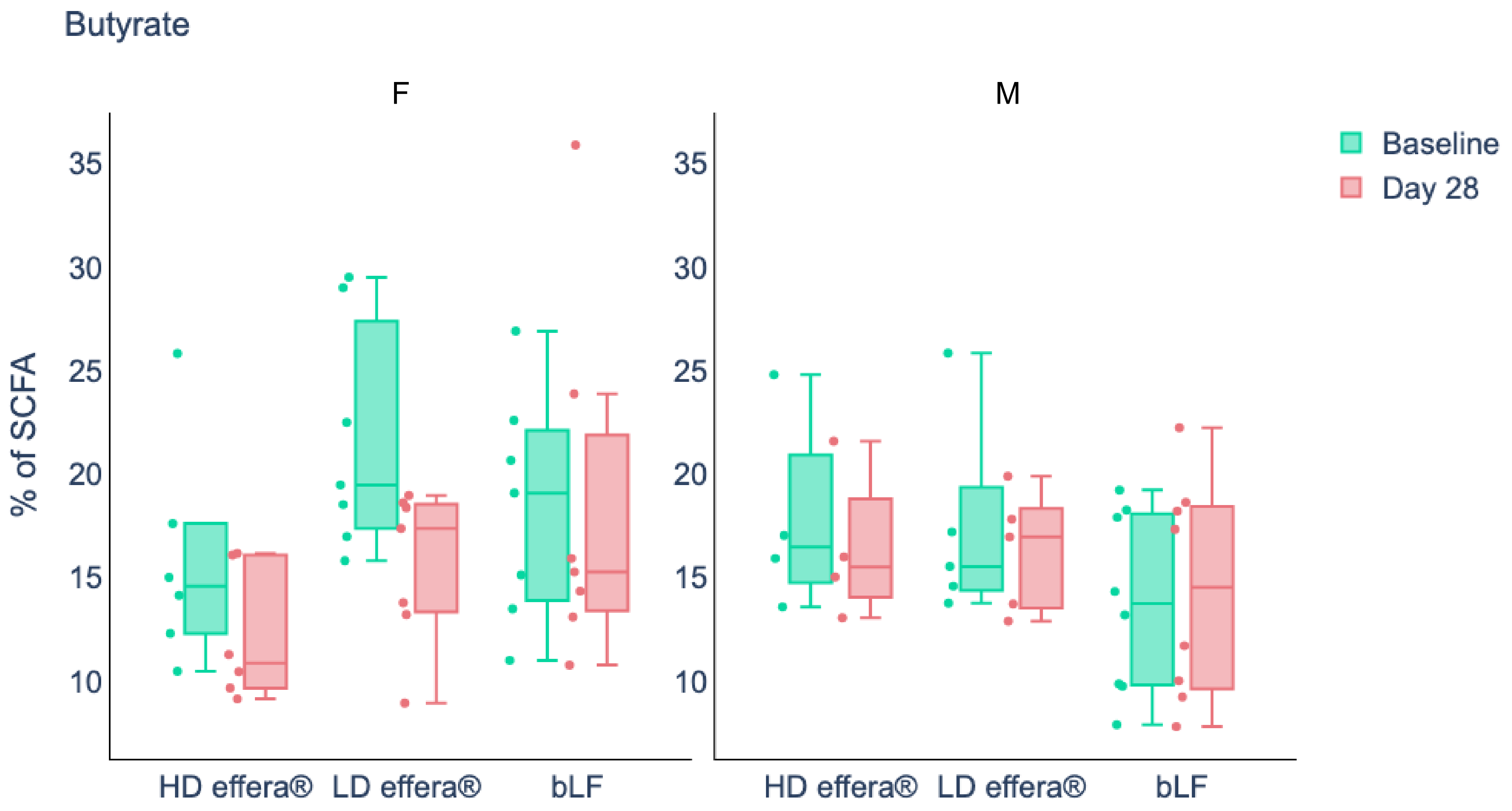


**Supplementary Figure A6.** Proportions of A) acetate, B) propionate, and C) butyrate at baseline (Day 0 and Day 28) for each treatment group by sex. High-dose effera® (3.4 g/d) (n = 10; ; F:6 M:4); Low-dose effera® (0.34 g/d) (n = 12; F:7 M:5); Bovine Lactoferrin (3.4 g/d) (n = 15; F:7 M:8). Abbreviations: HD, high-dose effera®; LD, low-dose effera®; bLF, bovine lactoferrin; F, female; M, male.
